# A mega-analysis of low frequency resting-state measures in mood and psychosis-spectrum disorders

**DOI:** 10.1101/2025.08.15.25332894

**Authors:** Maya L. Foster, Milana Khaitova, Saloni Mehta, Jean Ye, Dustin Scheinost

## Abstract

**Objective:** Conduct a mega-analysis of two complementary measures of resting-state functional magnetic resonance imaging (rsfMRI) dynamics—amplitude of low-frequency fluctuation (ALFF) and low-frequency spectral entropy (lfSE)—in mood and psychosis-spectrum disorders to evaluate group differences and clinical symptom associations.

**Design:** ALFF and lfSE were calculated at the node-level by filtering data from 0.01 Hz to 0.08 Hz, regressing demographic variables, and harmonizing sites. Group differences were assessed using the Wilcoxon signed test. Symptom associations were evaluated with Spearman’s rho. Analyses were conducted at both whole-brain and network levels, with sensitivity analyses to evaluate the impact of frequency brands.

**Setting:** Four independent open-source case-control datasets with resting-state functional magnetic resonance imaging were used: the Center for Biomedical Research Excellence, the Human Connectome Project for Early Psychosis, the Strategic Research Program for Brain Sciences, and the UCLA Consortium for Neuropsychiatric Phenomics.

**Participants:** Included participants had a mood disorder (bipolar, dysthymia, or major depressive disorder, n=228, aged 38.31 ± 12.56 years), a psychosis-spectrum disorder (early psychosis, schizophrenia spectrum disorder, or mood disorder with psychotic symptoms, n=318, aged 29.8 ± 13.21 years), or a healthy control (n=535, aged 39.89 ± 15.3 years).

**Main outcomes and Measures:** To identify group differences and symptom associations in mood and psychosis-spectrum disorders using ALFF and lfSE.

**Results:** ALFF in psychosis-spectrum was significantly lower than mood disorders and controls (q’s<0.001) at the whole-brain and network levels. lfSE in controls was significantly lower than both psychosis-spectrum and mood disorders at the whole-brain and network levels (q’s<0.001). Whole-brain ALFF is positively associated with mood symptoms (rho=0.27, p<0.05). Whole-brain lfSE is negatively associated with positive (rho=-0.13, p<0.05) and mood (rho=-0.38, p<0.01) symptoms. A greater sensitivity of group differences and symptom associations to frequency ranges was observed in mood disorders. ALFF is sensitive to medication.

**Conclusions and Relevance:** Widespread, global differences in ALFF and lfSE underly psychosis-spectrum and mood disorders. lfSE may be applicable for wider use in fMRI. Differences in spectral measures of brain dynamics may represent shared and distinct markers of mental health.

**Key Points:** *Question:* How do the amplitude and complexity of low-frequency oscillations in fMRI signals associate with psychosis-spectrum and mood disorders?

*Findings:* Findings suggest that the amplitude and complexity of low-frequency oscillation are associated with mood and psychosis-spectrum disorders in a wide-spread and global manner. Complexity, which is well-studied in EEG but not fMRI— emerged as a promising measure for further research. Our results also support a frequency-based behavioral encoding system operating in the BOLD signal.

*Meaning:* Our findings indicate that mood and psychosis-spectrum disorders differ from healthy controls in a mechanistically distinct way, as well as from one another across the whole brain. They also suggest that differences in low-frequency oscillations may represent shared and distinct markers of mental health and may help in developing treatment strategies that target these disruptions.

## INTRODUCTION

Mood (e.g., major depressive disorder and bipolar disorder) and psychosis-spectrum (e.g., first-episode psychosis and schizophrenia) disorders cause significant disability globally^1–4^ and current treatment outcomes remain suboptimal. These disorders also have shared and unique genetic risks^5–11^. Thus, continuing to advance knowledge of their shared and unique mechanisms will be important for improving differential diagnosis^11,12^ and symptom management for affected individuals. Widely associated with cognition and mental health^13–16^, changes in brain dynamics may be linked to the pathophysiology of mood and psychosis-spectrum disorders. However, while applications of brain dynamics methods are becoming increasingly popular, there is a scarcity of work exploring their simultaneous application to mood and psychosis-spectrum disorders, raising questions of whether linked dynamics signatures are shared, distinct, or both.

Spectral measures succinctly characterize brain dynamics across frequency bands^17–19^. One widely used metric is the amplitude of low-frequency fluctuations (ALFF), which quantifies the signal strength in the 0.01-0.08 Hz range and is well-established in functional neuroimaging^20–24^. A comparable measure is low-frequency spectral entropy (lfSE)—a frequency-bounded derivative of spectral entropy that quantifies the complexity or unpredictability of fMRI signal fluctuations in the 0.01–0.08 Hz range. Spectral entropy has been broadly used in EEG^24^, but not in fMRI studies. ALFF and lfSE provide distinct, but complementary insights (amplitude versus complexity) into intrinsic brain dynamics that may help biologically differentiate disorders and mediate individual differences in symptom severity.

Prior work has identified ALFF differences across major functional networks in mood^25–27^ and psychosis-spectrum^28–30,27^ disorders. In schizophrenia, studies found reduced ALFF in the default mode (DMN), salience (SAL), sensorimotor, parietal, and visual networks compared to healthy controls^31^. Early-stage schizophrenia has been marked by increased ALFF in subcortical and visual networks and decreased ALFF in the DMN and parietal networks^31^. In contrast, chronic schizophrenia includes widespread increases in ALFF across frontal, SAL, temporal, and limbic networks, alongside reductions in sensorimotor, parietal, and occipital areas^31^. In depression, studies report increased ALFF in several regions—including lateralized subcortical areas, occipital lobe, limbic areas, frontoparietal network (FPN), and the DMN— compared to matched healthy controls^26,32–35^. Decreased ALFF is also observed in somatosensory cortex^25,36,32^, limbic areas, and cerebellum^37,38^ for major depressive disorder (MDD) compared to controls. In bipolar disorder, increased ALFF in the right caudate and putamen^39^, bilateral insula, medial prefrontal cortex, and decreased ALFF in the left cerebellum^40^ have been observed. In contrast, lfSE has not been studied in mood and psychosis-spectrum disorders using fMRI. Moreover, to our knowledge no previous work has comprehensively contrasted ALFF with lfSE in mood and psychosis-spectrum disorders in one study.

Mega-analyses (e.g., a method that pools together the raw, individual level data from multiple studies), are a promising approach for finding shared and distinct biological indices across disorders. By pooling data from several smaller studies, mega-analyses maximize population variance, increase generalizability, and improve statistical power^41,42^. ALFF and lfSE are ideal for these analyses because they are computationally tractable in large samples, easy to pool across diverse datasets due to their conceptual simplicity, and neurobiologically interpretable at different spatial scales (i.e., regional, network, or whole brain levels). By better accounting for psychiatric heterogeneity, mega-analyses analyses can support the development of treatments that target shared and distinct clinical mechanisms^43–45^.

Here, we perform a mega-analysis in over 1000 individuals to compare ALFF and lfSE in individuals with mood disorders, individuals with psychosis-spectrum disorders and healthy controls. A secondary objective was to assess the utility of lfSE compared to ALFF. Our study expands on previous empirical findings by adding a complexity measure (i.e., lfSE)^46^, using a large-scale sample size (n=1081), and comparing measures across a wider range of symptoms. We also assess their association with relevant symptom measures. Based on findings from previous works, we hypothesized that ALFF and lfSE case-control group differences will be in the FPNl^47,48^, SAL^47^, and DMN^47,49^.

## METHODS

### Participants

We used four publicly available resting-state fMRI datasets (see Table S1 for participant demographics), the Center for Biomedical Research Excellence^50^ (COBRE, n=99), the Human Connectome Project Early Psychosis^51^ (HCP-EP, n=169), the Strategic Research Program for Brain Sciences^52^ (SRPBS, n=609) Multi-disorder Connectivity Dataset, and the University of California Los Angeles (UCLA) Consortium for Neuropsychiatric Phenomics^53^ (CNP, n=204). The sample included 535 healthy controls (HCs), 228 mood disorder (major depressive disorder, bipolar disorder, and dysthymia), and 318 psychosis-spectrum disorder (early psychosis and schizophrenia spectrum disorder) participants. All patients were diagnosed according to the DSM-5^54^. Demographics, symptom scores, medication information, and diagnosis breakdowns are summarized in the Supplement methods and tables S1, S5, and S6.

### Symptom Measures

Psychosis symptoms were assessed with the Positive and Negative Syndrome Scale^55^ (PANSS), which measures the presence and severity of positive, negative, and general psychopathology for an individual within the past week. The scale is widely used in clinical psychosis studies^56^ and has demonstrated reliability in assessing psychopathology of schizophrenia populations^57^. 230 participants had PANSS scores (HCP-EP=107, SRPBS=123). Mood symptoms in individuals (n=62) with a mood disorder (SRPBS=39) and health controls (SRPBS=23) were assessed with the Beck’s Depression Inventory^58^ (BDI-II). The BDI-II measures hallmark symptoms of depression and has high internal consistency^58,59^.

### Data Preprocessing

The acquisition and imaging parameters for the datasets are detailed elsewhere^50,51,60,52^. However, an abridged version is available in the Supplement (see Table S2). Images were motion and slice time corrected using SPM12^61^. BioImage Suite was used to perform mean white matter regression, cerebral spinal fluid regression, and gray matter time, removing linear trends, and low-pass filtering^62^. The Shen-268 atlas^61^ was warped from Montreal Neurological Institute (MNI) space into single-subject space. Next, the mean-time course of each node was calculated by averaging its constituent voxels’ time-series data.

### Quality Control

Participants were excluded if they had an average frame-to-frame displacement exceeding 0.2 mm (n=28), insufficient quality of linear or nonlinear registration (n=16), had >100 missing nodes (n=10) or had less than 75% coverage of relevant nodes for a given network or whole-brain analysis (n=15, see Supplement for more details on data preservation process). This left a final sample of 1081 subjects for further analysis: 823 for whole-brain analyses (averaging across all nodes) and varying sample sizes for network analyses (Table S3-4).

### ALFF Calculation and Analysis

First, the power spectral density of each node’s time series was calculated using periodogram function from the SciPy ^63^ signal processing API in Python 3.9. This calculation entailed converting each participant’s node time-series into the frequency domain via the fast Fourier transform (FFT). Then, the power spectral density was integrated over the low-frequency range (0.01 Hz ≤ frequency ≤ 0.08 Hz) using SciPy’s trapezoid integration method to calculate total power. Next, we took the square root of that operation to quantify ALFF.

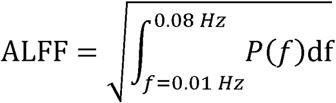

ALFF in this study is considered a proxy for the intensity of neural-associated brain activity or the energy content of the low-frequency band. A high ALFF signifies greater oscillation intensity while a low ALFF signifies lower oscillation intensity in the low-frequency range. ALFF was assessed group-wise at the whole-brain by averaging all nodes and network-levels using 10 canonical networks from the Shen 268 atlas.

### Low-frequency Spectral Entropy Calculation and Analysis

lfSE was also calculated at the node-level. Individual node time-series underwent a band-pass filter in the low-frequency range (0.01 Hz ≤ frequency ≤ 0.08 Hz) using SciPy’s butter and filtfilt functions (order = 2)^21^. The signal was transformed into the frequency domain using FFT, followed by power spectral density calculation and normalization to a probability density function. Then, lfSE was calculated as the Shannon entropy of the power spectral density was calculated using AntroPy^64^ package in Python 3.

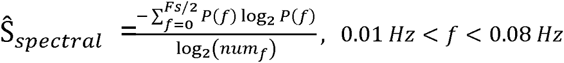

As a complexity measure, high lfSE (flatter power spectral density spectrum) represents low complexity while low lfSE (power spectral density distribution with more peaks) represents high complexity. lfSE was assessed group-wise at the whole-brain by averaging all nodes and network-levels using 10 canonical networks from the Shen 268 atlas.

### Statistical Analyses

Statistical analyses were constrained to the whole-brain and network levels to capture macroscale brain dynamics. Broader scale such as these have greater power than granular levels of inference^65^. Group comparisons of each low-frequency brain measure were evaluated at p<0.05 using the Wilcoxon two-sided rank sum test while correcting for multiple comparisons with false discovery rate (FDR). We controlled for self-reported age, motion, and sex as variables of non-interest. First, these factors were regressed^66^ using a generalized linear model on ALFF and lfSE values at the node level. Second, we applied the *neuroComba*t^67^ R package to harmonize data across sites. For empirical ranges, please refer to table S5). Where relevant, we report corrected (q) and uncorrected (p) results for transparency but only interpret significant corrected results (q<0.05).

### Secondary analyses

We correlated symptoms scores (PANSS and BDI-II) and covariate-regressed ALFF and lfSE values using Spearman’s correlation at the whole-brain and network levels. Participants for the PANSS analyses came from multiple sites and underwent site harmonization using *neuroCombat*^67^. Participants for the BDI-II analyses came from one site, so harmonization was not applied. Network-level associations were considered statistically significant at q<0.05 after FDR correction.

Additionally, we evaluated similarity between the ALFF and lfSE by correlating these values across participants (p<0.05) using Spearman’s rho at the whole-brain and network levels. We also calculated these similarities independently for each group. These correlations were Fisher transformed and compared across groups using a two-sample z-test.

### Sensitivity Analysis

We used different frequency cutoffs (0.01-0.045 Hz and 0.035-0.09 Hz) to assess if finer spectral divisions were driving the identified trends and associations^68^. We also examined the effect of medication exposure by including current medication status (binary: 1=on, 0=off) as an additional covariate.

## RESULTS

### Demographic differences between clinical groups

Significant group differences were found for self-reported sex (χ²=18.081, df = 2, p<0.001), age (W=75.48, df=2, p<0.001) and average frame displacement (W= 8.599, df=2, p=0.014). Participants with psychosis-spectrum (29.8 ± 13.21 years) were significantly younger than mood disorders (38.31 ± 12.56 years) and controls (39.89 ± 15.3 years). Participants with mood disorders also had significantly higher motion frame displacement (0.087 ± 0.045 mm) compared to matched HCs (0.083 ± 0.046 mm) and those with psychosis-spectrum disorder (0.075 ± 0.037 mm).

### Group differences in ALFF

At the whole-brain level, mood disorders exhibited the greatest ALFF, followed by controls and then psychosis-spectrum (Figure 2A, Table 1). Psychosis-spectrum ALFF values were significantly lower than both mood disorders and controls (p’s<0.001). No differences in whole-brain ALFF were detected between mood disorders and controls. Similar differences were observed at the network level, where all networks showed significant (q’s<0.05, FDR-corrected) differences when comparing psychosis-spectrum to mood disorders or health controls (Table 1). Results were similar when potential outliers were removed (Table S8) and when removing psychosis-spectrum patients with known affective disorders (Table S9).

**Figure 1.**
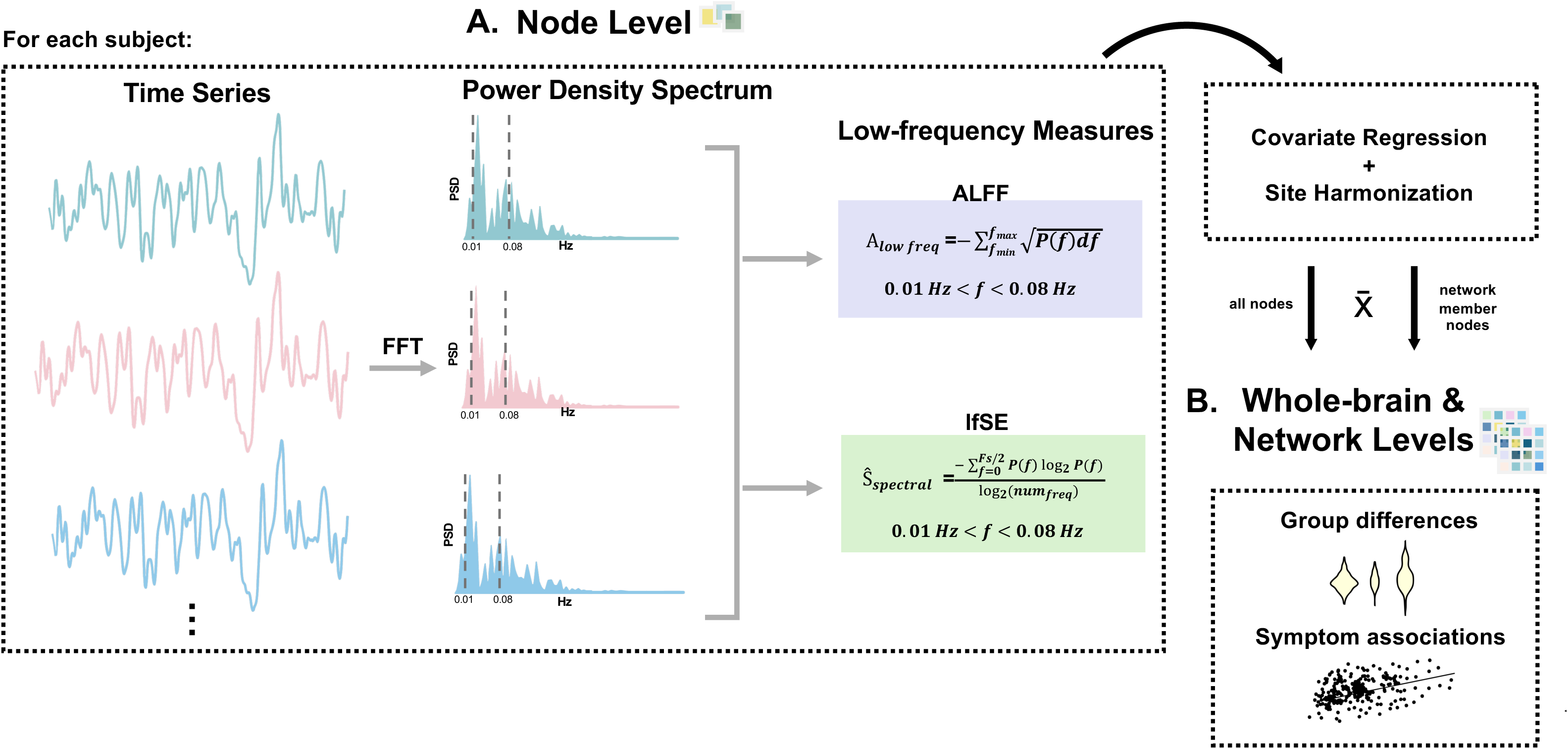
Schematic of analysis pipeline at node and network levels. This figure illustrates the study design and measurement calculation process. Four large-scale datasets were used in this study. A) First, the power spectrums were taken from preprocessed node-wise data, and the two low-frequency measures were calculated. Then, covariate regression of nuisance variables such as age, mean frame displacement, and sex were regressed using a generalized linear model. Where relevant, harmonization was performed to account for site effects. Then, the appropriate means were calculated for whole-brain (average over all nodes) and network (average over all member network nodes were taken for each of the 10 functional networks) analyses. B) Finally, group differences and symptom associations at whole-brain and network levels were calculated. FFT=Fast Fourier transform, ALFF=Amplitude of Low-Frequency Fluctuation, lfSE=low-frequency spectral entropy. x =mean.

**Figure 2.**
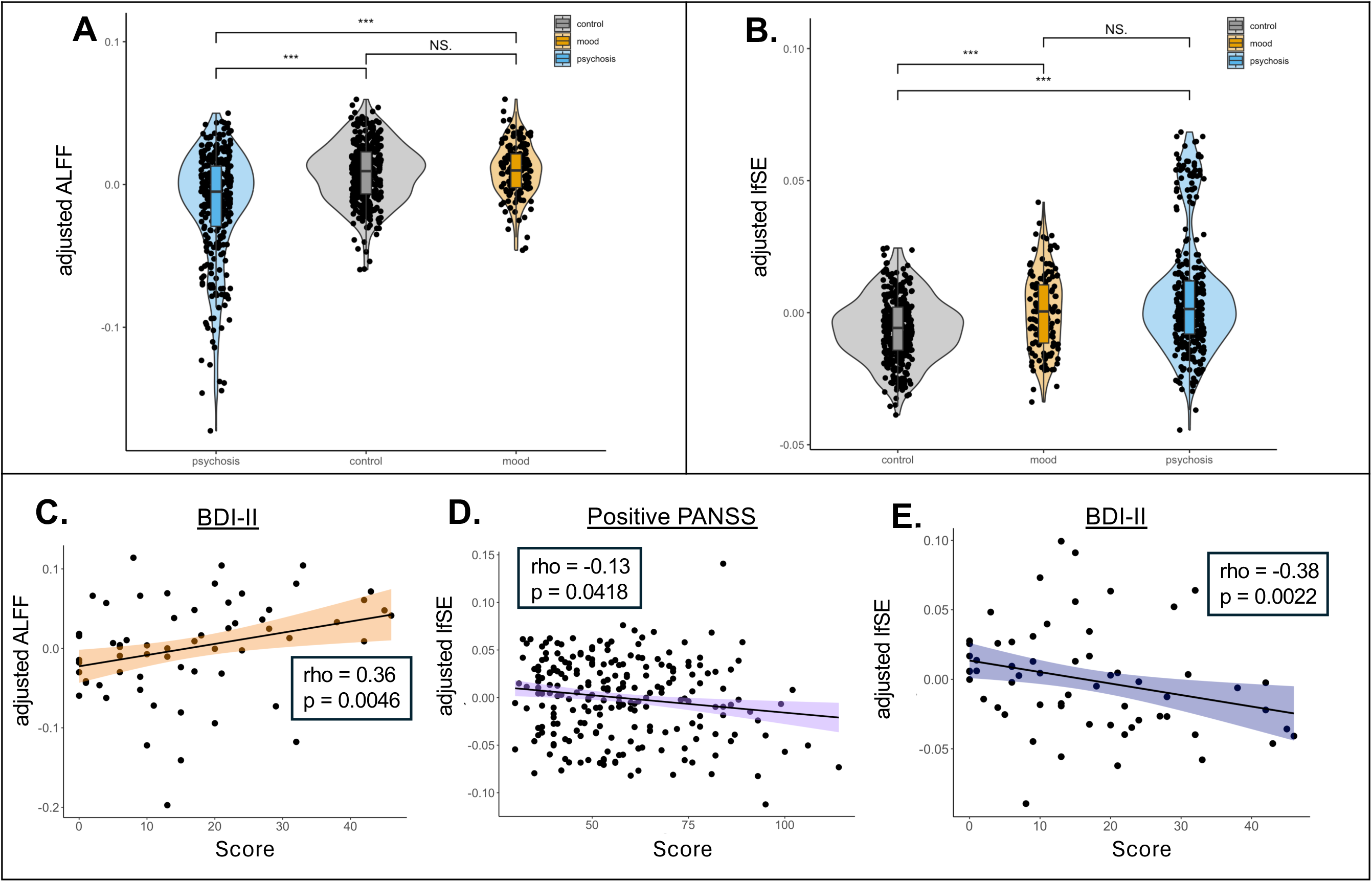
Group Differences at Whole-brain Level and Symptom Associations. Panels A and B feature a violin plot of the results for ALFF and lfSE respectively. Dots represent the participants (n=823; controls=366, mood=139, psychosis=318), colors represent the groups (controls, mood, psychosis). Lines of fit are based on linear model (for visualization). Correlations are reported as Spearman’s ρ. N.S. = not significant. ***=p<0.001. Panels C-E depict significant ALFF and lfSE symptom associations. Dots represent participants with a symptom score. A) Psychosis disorders had significantly lower ALFF than controls and mood disorders. B) Controls had significantly lower lfSE than mood and psychosis disorders. C) Significant BDI-II symptom associations with whole-brain ALFF (n=62). D) Significant positive PANSS symptom associations with whole-brain lfSE (n=230). E) Significant BDI-II symptom associations with whole-brain lfSE (n=62).

**Table 1.** Amplitude of low-frequency (ALFF, 0.01-0.08 Hz) value distributions by whole-brain, network and clinical group. Reported values have had covariates linearly regressed and undergone site harmonization. n=sample size. Bolded table entries represent significant comparisons after FDR correction.

| Scale | n | Clinical Group |  |  | Wilcoxon Rank Pairwise p |  |  |
| --- | --- | --- | --- | --- | --- | --- | --- |
|  |  | Control | Mood | Psychosis | C vs M | C vs P | M vs P |
| Whole-brain | 823 | 0.008 ±<br>0.021 | 0.01 ±<br>0.019 | -0.014 ±<br>0.039 | p = 0.399<br>q = 0.399 | **p = 5.88e-15**<br>**q = 1.77e-14** | **p = 1.83e-11**<br>**q = 2.74e-11** |
| Medial Frontal | 846 | 0.008 ±<br>0.025 | 0.011 ±<br>0.025 | -0.014 ±<br>0.041 | p = 0.149<br>q = 0.149 | **p = 4.28e-13**<br>**q = 1.28e-12** | **p = 2.68e-11**<br>**q = 4.02e-11** |
| Frontoparietal | 852 | 0.007 ±<br>0.022 | 0.01 ±<br>0.022 | -0.013 ±<br>0.04 | p = 0.146<br>q = 0.146 | **p = 5.49e-12**<br>**q = 1.65e-11** | **p = 7.03e-11**<br>**q = 1.05e-10** |
| Default Mode | 882 | 0.007 ±<br>0.026 | 0.011 ±<br>0.025 | -0.015 ±<br>0.045 | p = 0.0889<br>q = 0.0889 | **p = 3.77e-11**<br>**q = 1.13e-10** | **p = 9.94e-11**<br>**q = 1.49e-10** |
| Motor | 881 | 0.008 ±<br>0.025 | 0.011 ±<br>0.024 | -0.015 ±<br>0.042 | p = 0.212<br>q = 0.212 | **p = 2.34e-14**<br>**q = 7.01e-14** | **p = 4.57e-12**<br>**q = 6.86e-12** |
| Visual I | 1081 | 0.004 ±<br>0.028 | 0.008 ±<br>0.029 | -0.013 ±<br>0.043 | p = 0.0671<br>q = 0.0671 | **p = 3.63e-08**<br>**q = 5.44e-08** | **p = 9.89e-09**<br>**q = 2.97e-08** |
| Visual II | 953 | 0.006 ±<br>0.03 | 0.008 ±<br>0.031 | -0.013 ±<br>0.045 | p = 0.636<br>q = 0.636 | **p = 1.87e-09**<br>**q = 5.6e-09** | **p = 1.74e-07**<br>**q = 2.61e-07** |
| Visual Association | 906 | 0.008 ±<br>0.025 | 0.011 ±<br>0.029 | -0.016 ±<br>0.044 | p = 0.142<br>q = 0.142 | **p = 5.94e-14**<br>**q = 1.78e-13** | **p = 9.27e-12**<br>**q = 1.39e-11** |
| Salience | 936 | 0.006 ±<br>0.024 | 0.01 ±<br>0.025 | -0.014 ±<br>0.039 | p = 0.0987<br>q = 0.0987 | **p = 2.48e-12**<br>**q = 3.72e-12** | **p = 2.14e-12**<br>**q = 3.72e-12** |
| Subcortical | 906 | 0.007 ±<br>0.026 | 0.012 ±<br>0.028 | -0.016 ±<br>0.041 | p = 0.212<br>q = 0.212 | **p = 2.03e-14**<br>**q = 6.09e-14** | **p = 1.79e-12**<br>**q = 2.68e-12** |
| Cerebellar | 823 | 0.011 ±<br>0.033 | 0.012 ±<br>0.029 | -0.017 ±<br>0.053 | p = 0.998<br>q = 0.998 | **p = 6.45e-13**<br>**q = 1.93e-12** | **p = 1.17e-08**<br>**q = 1.75e-08** |

### Group differences in lfSE

At the whole-brain level, psychosis-spectrum had the highest lfSE, followed by mood disorders and then HCs (Figure 2B). Controls were significantly lower than both psychosis-spectrum and mood disorders at the whole-brain (p’s<0.001). No differences in whole-brain lfSE were detected between psychosis-spectrum and mood disorders. At the network level, all networks exhibited significant differences between health controls and psychosis-spectrum (q’s<0.05, FDR-corrected). The medial frontal, FPN, motor, subcortical, and cerebellar networks exhibited significant differences between HCs and mood disorders (q’s<0.05, FDR-corrected). While no differences between psychosis-spectrum and mood disorders existed at the whole-brain level, every network (other than the cerebellar network) exhibited significant between-group differences (q’s<0.05, FDR-corrected), likely due to the increases sample size used in the network analyses (Table 2). Results were similar when removing psychosis-spectrum patients with affective disorders (Table S10).

**Table 2.** Low-frequency spectral entropy (lfSE, 0.01-0.08 Hz) value distributions by whole-brain, network and clinical group. Reported values have had covariates linearly regressed and undergone site harmonization. n=sample size. Bolded table entries represent significant comparisons after FDR correction.

| Scale | n | Clinical Group |  |  | Wilcoxon Rank Pairwise p |  |  |
| --- | --- | --- | --- | --- | --- | --- | --- |
|  |  | Control | Mood | Psychosis | C vs M | C vs P | M vs P |
| Whole-brain | 823 | -0.006 ± 0.012 | 0 ± 0.015 | 0.007 ± 0.023 | **p = 1.52e-05**<br>**q = 2.28e-05** | **p = 2.5e-13**<br>**q = 7.49e-13** | p = 0.141<br>q = 0.141 |
| Medial Frontal | 846 | -0.006 ± 0.014 | -0.002 ± 0.015 | 0.008 ± 0.028 | **p = 0.00509**<br>**q = 0.00706** | **p = 8.25e-13**<br>**q = 2.48e-12** | **p = 0.00706**<br>**q = 0.00706** |
| Frontoparietal | 853 | -0.005 ± 0.014 | -0.001 ± 0.019 | 0.007 ± 0.025 | **p = 0.00107**<br>**q = 0.00161** | **p = 2.39e-10**<br>**q = 7.18e-10** | **p = 0.0463**<br>**q = 0.0463** |
| Default Mode | 882 | -0.004 ± 0.016 | -0.004 ± 0.019 | 0.007 ± 0.03 | p = 0.218<br>q = 0.218 | **p = 7.27e-08**<br>**q = 2.18e-07** | **p = 0.00191**<br>**q = 0.00287** |
| Motor | 881 | -0.006 ± 0.014 | -0.003 ± 0.019 | 0.008 ± 0.028 | **p = 0.00545**<br>**q = 0.00817** | **p = 4.66e-11**<br>**q = 1.4e-10** | **p = 0.00848**<br>**q = 0.00848** |
| Visual I | 1081 | -0.002 ± 0.017 | -0.009 ± 0.03 | 0.009 ± 0.026 | p = 0.0793<br>q = 0.0793 | **p = 1.95e-09**<br>**q = 3.37e-09** | **p = 2.25e-09**<br>**q = 3.37e-09** |
| Visual II | 954 | -0.003 ± 0.018 | -0.004 ± 0.027 | 0.007 ± 0.027 | p = 0.164<br>q = 0.164 | **p = 1.41e-08**<br>**q = 4.24e-08** | **p = 0.000741**<br>**q = 0.00111** |
| Visual Association | 906 | -0.005 ± 0.016 | -0.006 ± 0.027 | 0.01 ± 0.028 | p = 0.123<br>q = 0.123 | **p = 7.11e-13**<br>**q = 2.13e-12** | **p = 8.02e-05**<br>**q = 0.00012** |
| Saliency | 846 | -0.005 ± 0.013 | -0.004 ± 0.021 | 0.009 ± 0.027 | p = 0.0632<br>q = 0.0632 | **p = 4.85e-10**<br>**q = 1.46e-09** | **p = 0.000724**<br>**q = 0.00109** |
| Subcortical | 853 | -0.005 ± 0.013 | -0.005 ± 0.025 | 0.009 ± 0.026 | **p = 0.00158**<br>**q = 0.00237** | **p = 4.02e-12**<br>**q = 1.2e-11** | **p = 0.0309**<br>**q = 0.0309** |
| Cerebellar | 882 | -0.007 ± 0.019 | 0.002 ± 0.02 | 0.007 ± 0.034 | **p = 5.31e-06**<br>**q = 7.96e-06** | **p = 2.65e-08**<br>**q = 7.96e-08** | p = 0.641<br>q = 0.641 |

The psychosis-spectrum lfSE distribution was bimodal. Since the second (higher) peak in the distribution may influence the results, we repeated the analyses after removing these high-leverage points (see Supplemental Methods, Figure S4). Results were similar (Table S11); however, mood disorders now showed the highest lfSE, and only visual networks remained significantly different between psychosis-spectrum and mood disorders (q’s<0.05, FDR-corrected).

### Associations with psychosis and mood symptoms

Whole-brain ALFF is positively associated with BDI-II scores (rho=0.36, p= 0.0046; Figure 2C, Table S12) but had no association with PANSS scores (Table S13). Additionally, whole-brain lfSE is negatively associated with positive PANSS (rho=-0.13, p=0.0418, Table S14) and BDI-II scores (rho=-0.38, p=0.002; see Figure 2D-E, Table S15). No other significant associations with ALFF or lfSE at the whole-brain or network-level were observed (Tables S12-15).

### Correlation of ALFF and lfSE

At all scales of analysis, ALFF and lfSE were negatively correlated, such that individuals with higher ALFF values had lower lfSE values (rho’s = -0.39 to -0.22, Table S16-17). This relationship was also observed in our group comparison, where ALFF was consistently greater in mood disorder compared to psychosis-spectrum across scales and lfSE was consistently greater in psychosis-spectrum compared to mood disorder across scales.

### Sensitivity analyses of finer-scale frequency ranges

Results at the finer-scale frequency ranges were broadly similar to the main results, with wide-spread differences observed between groups (Tables S18-25). However, results involving the mood disorder group exhibited the most changes in finer-scale frequency ranges. For example, the lfSE of the DMN was significantly different between controls and mood disorders at 0.035-0.090 Hz, but not 0.01-0.045 Hz or the full frequency range. Additionally, whole-brain ALFF was positively associated with BDI-II scores at 0.01-0.045 Hz but negatively associated at 0.035-0.09 Hz (Table S22).

### Medication differentially impacts ALFF and lfSE

Results after controlling for medication were broadly similar for lfSE, but not ALFF (Table 3, Table S26). Only comparisons involving the mood disorder group were significant for ALFF. The ALFF comparisons between controls and psychosis spectrum were not significant.

**Table 3.** Effect of Medication Exposure on ALFF Group Differences. A subsample of the patient group had known exposure to a psychiatric medication and a sensitivity analysiswas performed on this subsample. Mood-psychosis effects remained consistent with analyses that did not consider medication exposure. However, case-control differences for mood disorders became significant, while those for psychosis-spectrum disorders became insignificant. Motor and visual I networks were analyzed using the mean only harmonization algorithm (see Supplementary methods).

| Scale | n | Clinical Groups (median +/- std) |  |  | Wilcoxon Rank Pairwise p |  |  |
| --- | --- | --- | --- | --- | --- | --- | --- |
|  |  | C | M | P | C vs M | C vs P | M vs P |
| Whole-brain | 554<br>(C=232<br>M=105<br>P=217) | -0.004 ±<br>0.021 | 0.026 ±<br>0.03 | -0.008 ±<br>0.039 | **p = 2.31e-18**<br>**q = 6.93e-18** | p = 0.937<br>q = 0.937 | **p = 1.56e-13**<br>**q = 2.34e-13** |
| Medial Frontal | 554<br>(C=232<br>M=105<br>P=217) | -0.004 ±<br>0.025 | 0.025 ±<br>0.032 | -0.008 ±<br>0.038 | **p = 7.97e-15**<br>**q = 2.39e-14** | p = 0.788<br>q = 0.788 | **p = 1.91e-12**<br>**q = 2.86e-12** |
| Frontoparietal | 554<br>(C=232<br>M=105<br>P=217) | -0.005 ±<br>0.026 | 0.022 ±<br>0.026 | -0.005 ±<br>0.038 | **p = 2.82e-16**<br>**q = 8.45e-16** | p = 0.539<br>q = 0.539 | **p = 1.98e-11**<br>**q = 2.97e-11** |
| Default Mode | 554<br>(C=232<br>M=105<br>P=217) | -0.004 ±<br>0.026 | 0.026 ±<br>0.034 | -0.008 ±<br>0.042 | **p = 1.52e-14**<br>**q = 4.56e-14** | p = 0.946<br>q = 0.946 | **p = 6.04e-12**<br>**q = 9.05e-12** |
| Motor | 555<br>(C=232<br>M=106<br>P=217) | -0.004 ±<br>0.024 | 0.025 ±<br>0.032 | -0.008 ±<br>0.039 | **p = 3.87e-15**<br>**q = 1.16e-14** | p = 0.755<br>q = 0.755 | **p = 1.88e-12**<br>**q = 2.82e-12** |
| Visual I | 558<br>(C=232<br>M=109,<br>P=217) | -0.004 ±<br>0.027 | 0.019 ±<br>0.031 | -0.006 ±<br>0.04 | **p = 4.76e-11**<br>**q = 1.43e-10** | p = 0.92<br>q = 0.92 | **p = 6.96e-08**<br>**q = 1.04e-07** |
| Visual II | 556<br>(C=232<br>M=107<br>P=217) | -0.004 ±<br>0.028 | 0.026 ±<br>0.039 | -0.008 ±<br>0.043 | **p = 7.82e-14**<br>**q = 2.35e-13** | p = 0.645<br>q = 0.645 | **p = 1.12e-10**<br>**q = 1.68e-10** |
| Visual Association | 555<br>(C=232<br>M=106<br>P=217) | -0.004 ±<br>0.025 | 0.025 ±<br>0.034 | -0.008 ±<br>0.041 | **p = 3.55e-15**<br>**q = 1.07e-14** | p = 0.722<br>q = 0.722 | **p = 1.55e-12**<br>**q = 2.33e-12** |
| Salience | 555<br>(C=232<br>M=106<br>P=217) | -0.004 ±<br>0.023 | 0.024 ±<br>0.031 | -0.007 ±<br>0.038 | **p = 2.49e-15**<br>**q = 7.47e-15** | p = 0.871<br>q = 0.871 | **p = 1.21e-11**<br>**q = 1.81e-11** |
| Subcortical | 554<br>(C=232<br>M=105<br>P=217) | -0.004 ±<br>0.024 | 0.024 ±<br>0.032 | -0.008 ±<br>0.04 | **p = 4.28e-13**<br>**q = 1.28e-12** | p = 0.416<br>q = 0.416 | **p = 1.29e-10**<br>**q = 1.94e-10** |
| Cerebellar | 554<br>(C=232<br>M=105<br>P=217) | -0.005 ±<br>0.03 | 0.03 ±<br>0.042 | -0.009 ±<br>0.055 | **p = 5.36e-14**<br>**q = 1.61e-13** | p = 0.643<br>q = 0.643 | **p = 2e-09**<br>**q = 3e-09** |

## DISCUSSION

In this study, we conducted a mega-analysis using two spectral measures— oscillation intensity (ALFF) and complexity (lfSE) —to compare mood and psychosis-spectrum disorders. Our results highlight extensive differences between these two clinical conditions that span multiple dimensions of brain dynamics.

Notably, ALFF and lfSE uniquely differentiated clinical groups, providing distinct insights into the biological underpinnings of mood and psychosis-spectrum disorders. Psychosis-spectrum participants exhibited larger, widespread differences in lfSE compared to mood disorder participants and controls, while mood disorder participants exhibited more focal differences in lfSE compared to controls. Also, in line with several studies ^25,26,32–38^, we observed significant differences between mood disorder participants and controls in ALFF after controlling for medication. In some cases, ALFF and lfSE results even converged. For example, despite our initial hypotheses, there is widespread overlap in the functional networks that had significant group differences between mood and psychosis-spectrum disorders.

Our sensitivity analyses revealed that ALFF results in mood disorders vary by frequency band. In the narrower range of 0.035 to 0.090 Hz, larger group differences and significant associations with mood symptoms were discovered. Moreover, there was a reversal in the sign of significant mood symptom associations in the two finer-scale frequency bands (0.01-0.045 Hz and 0.035-0.090 Hz). Aligned with these findings, a previous study showed that associations between depressive symptoms and spectral measures in the subgenual gyrus differ between frequency bands^69^. These diverging associations are evidence of the functional specificity of frequency bands in the blood-oxygen level dependent (BOLD) signal^70–72^, a phenomena that has largely been attributed to EEG studies of mental health^73^. As such, the notion that different frequency bands reflect physiology occurring at a range of intrinsic time scales^73–76^ is likely a universal principle of brain function. Under this consideration, the finer bands represented in our work comprise multiple physiological processes that contribute to mood disorder pathology. If too broad a frequency range is adopted, interactive effects^69^ can dominate and obscure significant results in a competitive or mutually constructive way^77^. Continued investigations of the functional relevance of specific frequency bands in mood disorders and other psychiatric conditions are needed in fMRI research.

Out of the two measures, lfSE performed the best in detecting significant group differences and symptom associations. It was also robust to common sources of variance, which is crucial for mega-analyses, as they often encompass real-world conditions (e.g., use of different scanners, participants with varying medication histories). lfSE may more effectively capture changes in brain dynamics because as a measure of complexity and irregularity^78^, it considers the entire energy distribution across a frequency band. Previous work employing similar methods in other imaging modalities have identified signal irregularity and high signal variance as key features of the psychosis-spectrum^79–82^. Our results, alongside broader literature findings, position lfSE—a standard measure in the EEG literature^24^ and related modalities^78,83–85^—as a valuable index for increased application in fMRI. Future research should apply the lfSE metric under different conditions and test its robustness against different covariates to fully assess its mechanistic insights and resilience to non-interest variance sources.

In contrast, we observed a change in the significance of two group difference pairs in ALFF analyses with and without medication (Table 3). This inconsistency may indicate that ALFF is more susceptible to medication effects, or more specifically, medication effects in mood and psychosis-spectrum disorders. Future studies should further probe ALFF’s sensitivity to medication in other psychiatric populations to confirm.

This study has several strengths. We combined distinct diagnoses into broader groupings (e.g., mood and psychosis-spectrum disorders), which may better reflect the dimensional nature of disorders and allow datasets with different diagnostic criteria to be combined in a mega-analysis. We also used two spectral measures, including lfSE (which is uncommon in fMRI analysis). It also has several limitations. First, participants were assumed to exclusively have mood or a psychosis-spectrum disorder. However, comorbidities are common, and individuals could belong in multiple groupings ^86^. Second, we focus on measures that compress brain dynamics into a single number. More complex summaries of dynamic measures^87^ may prove to be more sensitive and is a natural next step. Third, although we identify associations with symptoms scores, causality cannot be established.

Through a mega-analysis, we investigated group differences using spectral measures of brain dynamics in mood and psychosis-spectrum disorders. Our approach revealed distinct brain signal properties in addition to shared characteristics between the two patient groups. Future studies should continue to elucidate common denominators as well as diagnosis-, symptom-, and individual-specific brain deviations that build on top of disorder commonalities. Better characterization of shared brain features, and disorder specific deviations could pave the way towards plausible prevention and tailored treatment efforts.

## Data Availability

All data produced are available online.

https://openneuro.org/datasets/ds000030/versions/00016

https://www.humanconnectome.org/study/human-connectome-project-for-early-psychosis

http://fcon_1000.projects.nitrc.org/indi/retro/cobre.html

https://bicr-resource.atr.jp/srpbsopen/

## Data and Code Availability

Data are publicly available through the Center for Biomedical Research Excellence Phase I grant^50^, the Human Connectome Project Early Psychosis^51^, the Strategic Research Program for Brain Sciences^52^ and the UCLA Consortium for Neuropsychiatric Phenomics LA5c Study^88^. (https://openneuro.org/datasets/ds000030/versions/00016). Preprocessing scripts are available at https://www.nitrc.org/projects/bioimagesuite.

## Acknowledgements

We are grateful to the participants who dedicated their time and energy to participate in these large-scale data consortiums as well as the study team who collected and collated the data. We also would like to thank Dr. Michelle Hampson and Dr. Nicha Dvornek for initial input that guided the evolution of this research project and Dr. Margaret L. Westwater for critically insightful feedback on the manuscript.

## Author Contributions and Assistive Grammar Tools Acknowledgement

MF had full access to the data and takes on full responsibility for accuracy of the data analysis. MF conceived and designed the study. MF participated in the acquisition, preprocessing, analysis, and interpretation of data. MF and DS participated in drafting the manuscript. MF and DS performed statistical analyses. SM and JY provided critical review of the content and provided clinical interpretations of the analyses. MK participated in quality control of data. JY participated in acquisition and preprocessing of some of the data.

Portions of the writing in this manuscript were edited with artificial intelligence (AI)-based tools to improve clarity and grammar. The authors critically reviewed and edited all AI-assisted content to ensure accuracy and originality. No AI tools were used in the generation of scientific content, data analysis, or interpretation. All intellectual content, interpretations, and conclusions are our own.

## REFERENCES

1. Ayano G, Demelash S, yohannes Z, et al. Misdiagnosis, detection rate, and associated factors of severe psychiatric disorders in specialized psychiatry centers in Ethiopia. Ann Gen Psychiatry. 2021;20:10. doi:10.1186/s12991-021-00333-7

2. Calabrese J, Al Khalili Y. Psychosis. In: StatPearls. StatPearls Publishing; 2024. Accessed March 31, 2024. http://www.ncbi.nlm.nih.gov/books/NBK546579/

3. Sekhon S, Gupta V. Mood Disorder. In: StatPearls. StatPearls Publishing; 2025. Accessed February 14, 2025. http://www.ncbi.nlm.nih.gov/books/NBK558911/

4. Global, regional, and national burden of 12 mental disorders in 204 countries and territories, 1990–2019: a systematic analysis for the Global Burden of Disease Study 2019. The Lancet Psychiatry. 2022;9(2):137–150. doi:10.1016/S2215-0366(21)00395-3

5. International Schizophrenia Consortium, Purcell SM, Wray NR, et al. Common polygenic variation contributes to risk of schizophrenia and bipolar disorder. Nature. 2009;460(7256):748–752. doi:10.1038/nature08185

6. Identification of risk loci with shared effects on five major psychiatric disorders: a genome-wide analysis. The Lancet. 2013;381(9875):1371–1379. doi:10.1016/S0140-6736(12)62129-1

7. Ruderfer DM, Ripke S, McQuillin A, et al. Genomic dissection of bipolar disorder and schizophrenia including 28 subphenotypes. Cell. 2018;173(7):1705–1715.e16. doi:10.1016/j.cell.2018.05.046

8. Fusar-Poli P, Solmi M, Brondino N, et al. Transdiagnostic psychiatry: a systematic review. World Psychiatry. 2019;18(2):192–207. doi:10.1002/wps.20631

9. Bigdeli TB, Fanous AH, Li Y, et al. Genome-Wide Association Studies of Schizophrenia and Bipolar Disorder in a Diverse Cohort of US Veterans. Schizophr Bull. 2020;47(2):517–529. doi:10.1093/schbul/sbaa133

10. Xie C, Xiang S, Shen C, et al. A shared neural basis underlying psychiatric comorbidity. Nat Med. 2023;29(5):1232–1242. doi:10.1038/s41591-023-02317-4

11. Brand SJ, Möller M, Harvey BH. A Review of Biomarkers in Mood and Psychotic Disorders: A Dissection of Clinical vs. Preclinical Correlates. Curr Neuropharmacol. 2015;13(3):324–368. doi:10.2174/1570159X13666150307004545

12. Kas MJH, Penninx BWJH, Knudsen GM, et al. Precision psychiatry roadmap: towards a biology-informed framework for mental disorders. Mol Psychiatry. 2025;30(8):3846–3855. doi:10.1038/s41380-025-03070-5

13. Newson JJ, Thiagarajan TC. EEG Frequency Bands in Psychiatric Disorders: A Review of Resting State Studies. Front Hum Neurosci. 2019;12. doi:10.3389/fnhum.2018.00521

14. Raut RV, Snyder AZ, Mitra A, et al. Global waves synchronize the brain’s functional systems with fluctuating arousal. Sci Adv. 2021;7(30):eabf2709. doi:10.1126/sciadv.abf2709

15. Xu Y, Long X, Feng J, Gong P. Interacting spiral wave patterns underlie complex brain dynamics and are related to cognitive processing. Nat Hum Behav. 2023;7(7):1196–1215. doi:10.1038/s41562-023-01626-5

16. Foster M, Scheinost D. Brain states as wave-like motifs. Trends in Cognitive Sciences. Published online 2024. Accessed April 29, 2024. https://www.cell.com/trends/cognitive-sciences/abstract/S1364-6613(24)00057-3

17. Duff EP, Johnston LA, Xiong J, Fox PT, Mareels I, Egan GF. The power of spectral density analysis for mapping endogenous BOLD signal fluctuations. Human Brain Mapping. 2008;29(7):778–790. doi:10.1002/hbm.20601

18. Elble RJ, Ondo W. Tremor rating scales and laboratory tools for assessing tremor. J Neurol Sci. 2022;435:120202. doi:10.1016/j.jns.2022.120202

19. Zang Z, Qiao Y, Yan S, Lu J. Reliability and Validity of Power Spectrum Slope (PSS): A Metric for Measuring Resting-State Functional Magnetic Resonance Imaging Activity of Single Voxels. Front Neurosci. 2022;16:871609. doi:10.3389/fnins.2022.871609

20. Yang H, Long XY, Yang Y, et al. Amplitude of low frequency fluctuation within visual areas revealed by resting-state functional MRI. Neuroimage. 2007;36(1):144–152. doi:10.1016/j.neuroimage.2007.01.054

21. Zou QH, Zhu CZ, Yang Y, et al. An improved approach to detection of amplitude of low-frequency fluctuation (ALFF) for resting-state fMRI: Fractional ALFF. Journal of Neuroscience Methods. 2008;172(1):137–141. doi:10.1016/j.jneumeth.2008.04.012

22. Zuo XN, Di Martino A, Kelly C, et al. The Oscillating Brain: Complex and Reliable. Neuroimage. 2010;49(2):1432–1445. doi:10.1016/j.neuroimage.2009.09.037

23. Li MT, Zhang SX, Li X, et al. Amplitude of Low-Frequency Fluctuation in Multiple Frequency Bands in Tension-Type Headache Patients: A Resting-State Functional Magnetic Resonance Imaging Study. Front Neurosci. 2021;15. doi:10.3389/fnins.2021.742973

24. Yen C, Lin CL, Chiang MC. Exploring the Frontiers of Neuroimaging: A Review of Recent Advances in Understanding Brain Functioning and Disorders. Life. 2023;13(7):1472. doi:10.3390/life13071472

25. Wang L, Dai W, Su Y, et al. Amplitude of Low-Frequency Oscillations in First-Episode, Treatment-Naive Patients with Major Depressive Disorder: A Resting-State Functional MRI Study. PLOS ONE. 2012;7(10):e48658. doi:10.1371/journal.pone.0048658

26. Chen Q, Bi Y, Zhao X, et al. Regional amplitude abnormities in the major depressive disorder: A resting-state fMRI study and support vector machine analysis. Journal of Affective Disorders. 2022;308:1–9. doi:10.1016/j.jad.2022.03.079

27. Chen C, Zhang B, Qin X, et al. Altered resting-state brain activity of the superior parietal cortex and striatum in major depressive disorder and schizophrenia. Asian Journal of Psychiatry. 2024;102:104303. doi:10.1016/j.ajp.2024.104303

28. Turner JA, Damaraju E, Van Erp TGM, et al. A multi-site resting state fMRI study on the amplitude of low frequency fluctuations in schizophrenia. Front Neurosci. 2013;7. doi:10.3389/fnins.2013.00137

29. Wang P, Yang J, Yin Z, et al. Amplitude of low-frequency fluctuation (ALFF) may be associated with cognitive impairment in schizophrenia: a correlation study. BMC Psychiatry. 2019;19:30. doi:10.1186/s12888-018-1992-4

30. Li X, Liu Q, Chen Z, et al. Abnormalities of Regional Brain Activity in Patients With Schizophrenia: A Longitudinal Resting-State fMRI Study. Schizophrenia Bulletin. 2023;49(5):1336–1344. doi:10.1093/schbul/sbad054

31. Gong J, Wang J, Luo X, et al. Abnormalities of intrinsic regional brain activity in first-episode and chronic schizophrenia: a meta-analysis of resting-state functional MRI. J Psychiatry Neurosci. 2020;45(1):55–68. doi:10.1503/jpn.180245

32. Wu Z, Luo Q, Wu H, et al. Amplitude of Low-Frequency Oscillations in Major Depressive Disorder With Childhood Trauma. Front Psychiatry. 2021;11:596337. doi:10.3389/fpsyt.2020.596337

33. Zhang B, Qi S, Liu S, Liu X, Wei X, Ming D. Altered spontaneous neural activity in the precuneus, middle and superior frontal gyri, and hippocampus in college students with subclinical depression. BMC Psychiatry. 2021;21(1):280. doi:10.1186/s12888-021-03292-1

34. Zhang Z, Bo Q, Li F, et al. Increased ALFF and functional connectivity of the right striatum in bipolar disorder patients. Progress in Neuro-Psychopharmacology and Biological Psychiatry. 2021;111:110140. doi:10.1016/j.pnpbp.2020.110140

35. Luo G, Zhou J, Liu L, Song X, Peng M, Zhang X. Abnormal ReHo and ALFF values in drug-naïve depressed patients with suicidal ideation or attempts: Evidence from the REST-meta-MDD consortium. Progress in Neuro-Psychopharmacology and Biological Psychiatry. 2025;136:111210. doi:10.1016/j.pnpbp.2024.111210

36. Liu F, Guo W, Liu L, et al. Abnormal amplitude low-frequency oscillations in medication-naive, first-episode patients with major depressive disorder: a resting-state fMRI study. J Affect Disord. 2013;146(3):401–406. doi:10.1016/j.jad.2012.10.001

37. Liu J, Ren L, Womer FY, et al. Alterations in amplitude of low frequency fluctuation in treatment-naïve major depressive disorder measured with resting-state fMRI. Human Brain Mapping. 2014;35(10):4979–4988. doi:10.1002/hbm.22526

38. Zhou M, Hu X, Lu L, et al. Intrinsic cerebral activity at resting state in adults with major depressive disorder: A meta-analysis. Progress in Neuro-Psychopharmacology and Biological Psychiatry. 2017;75:157–164. doi:10.1016/j.pnpbp.2017.02.001

39. Zhang Z, Bo Q, Li F, et al. Increased ALFF and functional connectivity of the right striatum in bipolar disorder patients. Prog Neuropsychopharmacol Biol Psychiatry. 2021;111:110140. doi:10.1016/j.pnpbp.2020.110140

40. Gong J, Wang J, Qiu S, et al. Common and distinct patterns of intrinsic brain activity alterations in major depression and bipolar disorder: voxel-based meta-analysis. Transl Psychiatry. 2020;10(1):353. doi:10.1038/s41398-020-01036-5

41. Costafreda SG. Pooling fMRI data: meta-analysis, mega-analysis and multi-center studies. Front Neuroinform. 2009;3. doi:10.3389/neuro.11.033.2009

42. Bigdely-Shamlo N, Touryan J, Ojeda A, Kothe C, Mullen T, Robbins K. Automated EEG mega-analysis I: Spectral and amplitude characteristics across studies. NeuroImage. 2020;207:116361. doi:10.1016/j.neuroimage.2019.116361

43. Zambrano-Vazquez L, Levy HC, Belleau EL, et al. Using the research domain criteria framework to track domains of change in comorbid PTSD and SUD. Psychol Trauma. 2017;9(6):679–687. doi:10.1037/tra0000257

44. McQuaid RJ. Transdiagnostic biomarker approaches to mental health disorders: Consideration of symptom complexity, comorbidity and context. Brain Behav Immun Health. 2021;16:100303. doi:10.1016/j.bbih.2021.100303

45. Holmes AJ, Patrick LM. The Myth of Optimality in Clinical Neuroscience. Trends in Cognitive Sciences. 2018;22(3):241–257. doi:10.1016/j.tics.2017.12.006

46. Tenev A, Markovska-Simoska S, Müller A, Mishkovski I. Entropy, complexity, and spectral features of EEG signals in autism and typical development: a quantitative approach. Front Psychiatry. 2025;16. doi:10.3389/fpsyt.2025.1505297

47. Menon V. Large-scale brain networks and psychopathology: a unifying triple network model. Trends Cogn Sci. 2011;15(10):483–506. doi:10.1016/j.tics.2011.08.003

48. Foster ML, Ye J, Powers AR, Dvornek NC, Scheinost D. Connectome-based predictive modeling of early and chronic psychosis symptoms. Neuropsychopharmacol. Published online February 27, 2025:1–9. doi:10.1038/s41386-025-02064-9

49. Schilbach L, Hoffstaedter F, Müller V, et al. Transdiagnostic commonalities and differences in resting state functional connectivity of the default mode network in schizophrenia and major depression. NeuroImage: Clinical. 2016;10:326–335. doi:10.1016/j.nicl.2015.11.021

50. COBRE Phase 3 | The Mind Research Network (MRN). Accessed February 14, 2025. https://www.mrn.org/common/cobre-phase-3

51. Lewandowski KE, Bouix S, Ongur D, Shenton ME. Neuroprogression across the Early Course of Psychosis. J Psychiatr Brain Sci. 2020;5:e200002. doi:10.20900/jpbs.20200002

52. Tanaka SC, Yamashita A, Yahata N, et al. A multi-site, multi-disorder resting-state magnetic resonance image database. Sci Data. 2021;8(1):227. doi:10.1038/s41597-021-01004-8

53. Poldrack RA, Congdon E, Triplett W, et al. A phenome-wide examination of neural and cognitive function. Sci Data. 2016;3(1):160110. doi:10.1038/sdata.2016.110

54. American Psychiatric Association. Diagnostic and Statistical Manual of Mental Disorders. Fifth Edition. American Psychiatric Association; 2013. Accessed March 26, 2025. https://psychiatryonline.org/doi/book/10.1176/appi.books.9780890425596

55. Kay SR, Fiszbein A, Opler LA. The positive and negative syndrome scale (PANSS) for schizophrenia. Schizophr Bull. 1987;13(2):261–276. doi:10.1093/schbul/13.2.261

56. Shafer A, Dazzi F. Meta-analysis of the positive and Negative Syndrome Scale (PANSS) factor structure. Journal of Psychiatric Research. 2019;115:113–120. doi:10.1016/j.jpsychires.2019.05.008

57. Kay SR, Opler LA, Lindenmayer JP. Reliability and validity of the positive and negative syndrome scale for schizophrenics. Psychiatry Res. 1988;23(1):99–110. doi:10.1016/0165-1781(88)90038-8

58. Beck AT, Ward CH, Mendelson M, Mock J, Erbaugh J. An inventory for measuring depression. Arch Gen Psychiatry. 1961;4:561–571. doi:10.1001/archpsyc.1961.01710120031004

59. Beck AT, Steer RA, Carbin MG. Psychometric properties of the Beck Depression Inventory: Twenty-five years of evaluation. Clinical Psychology Review. 1988;8(1):77–100. doi:10.1016/0272-7358(88)90050-5

60. Gorgolewski KJ, Durnez J, Poldrack RA. Preprocessed Consortium for Neuropsychiatric Phenomics dataset. F1000Res. 2017;6:1262. doi:10.12688/f1000research.11964.2

61. Shen X, Tokoglu F, Papademetris X, Constable RT. Groupwise whole-brain parcellation from resting-state fMRI data for network node identification. Neuroimage. 2013;82:403–415. doi:10.1016/j.neuroimage.2013.05.081

62. Shen X, Finn ES, Scheinost D, et al. Using connectome-based predictive modeling to predict individual behavior from brain connectivity. Nat Protoc. 2017;12(3):506–518. doi:10.1038/nprot.2016.178

63. Virtanen P, Gommers R, Oliphant TE, et al. SciPy 1.0: fundamental algorithms for scientific computing in Python. Nat Methods. 2020;17(3):261–272. doi:10.1038/s41592-019-0686-2

64. antropy.app_entropy — antropy 0.1.9 documentation. Accessed July 24, 2025. https://raphaelvallat.com/antropy/build/html/generated/antropy.app_entropy.html

65. Noble S, Mejia AF, Zalesky A, Scheinost D. Improving power in functional magnetic resonance imaging by moving beyond cluster-level inference. Proceedings of the National Academy of Sciences. 2022;119(32):e2203020119. doi:10.1073/pnas.2203020119

66. Schöttner M, Bolton TAW, Patel J, Nahálka AT, Vieira S, Hagmann P. Exploring the latent structure of behavior using the Human Connectome Project’s data. Sci Rep. 2023;13(1):713. doi:10.1038/s41598-022-27101-1

67. Package neuroCombat details | Neuroconductor. Accessed August 25, 2025. https://neuroconductor.org/package/neuroCombat#citation

68. Zheng R, Chen Y, Jiang Y, et al. Dynamic Altered Amplitude of Low-Frequency Fluctuations in Patients With Major Depressive Disorder. Front Psychiatry. 2021;12. doi:10.3389/fpsyt.2021.683610

69. Song X, Hu X, Zhou S, et al. Association of specific frequency bands of functional MRI signal oscillations with motor symptoms and depression in Parkinson’s disease. Sci Rep. 2015;5(1):16376. doi:10.1038/srep16376

70. Caplan JB, Madsen JR, Raghavachari S, Kahana MJ. Distinct Patterns of Brain Oscillations Underlie Two Basic Parameters of Human Maze Learning. Journal of Neurophysiology. 2001;86(1):368–380. doi:10.1152/jn.2001.86.1.368

71. Ries A, Hollander M, Glim S, Meng C, Sorg C, Wohlschläger A. Frequency-Dependent Spatial Distribution of Functional Hubs in the Human Brain and Alterations in Major Depressive Disorder. Front Hum Neurosci. 2019;13:146. doi:10.3389/fnhum.2019.00146

72. Bonnefond M, Kastner S, Jensen O. Communication between Brain Areas Based on Nested Oscillations. eNeuro. 2017;4(2). doi:10.1523/ENEURO.0153-16.2017

73. Attar ET. Review of electroencephalography signals approaches for mental stress assessment. Neurosciences (Riyadh). 2022;27(4):209–215. doi:10.17712/nsj.2022.4.20220025

74. Buzsáki G, Draguhn A. Neuronal oscillations in cortical networks. Science. 2004;304(5679):1926-1929. doi:10.1126/science.1099745

75. Motivation, emotion, and their inhibitory control mirrored in brain oscillations - PubMed. Accessed August 7, 2025. https://pubmed.ncbi.nlm.nih.gov/17145079/

76. Liu ZZ, Qu HJ, Tian ZL, et al. Reproducibility of frequency-dependent low frequency fluctuations in reaction time over time and across tasks. PLOS ONE. 2017;12(9):e0184476. doi:10.1371/journal.pone.0184476

77. Guo X, Li J, Su Q, et al. Transcriptional correlates of frequency-dependent brain functional activity associated with symptom severity in degenerative cervical myelopathy. NeuroImage. 2023;284:120451. doi:10.1016/j.neuroimage.2023.120451

78. Helakari H, Kananen J, Huotari N, et al. Spectral entropy indicates electrophysiological and hemodynamic changes in drug-resistant epilepsy – A multimodal MREG study. NeuroImage: Clinical. 2019;22:101763. doi:10.1016/j.nicl.2019.101763

79. Yang GJ, Murray JD, Repovs G, et al. Altered global brain signal in schizophrenia. Proceedings of the National Academy of Sciences. 2014;111(20):7438–7443. doi:10.1073/pnas.1405289111

80. Bachiller A, Lubeiro A, Díez Á, et al. Decreased entropy modulation of EEG response to novelty and relevance in schizophrenia during a P300 task. Eur Arch Psychiatry Clin Neurosci. 2015;265(6):525–535. doi:10.1007/s00406-014-0525-5

81. Akar SA, Kara S, Latifoğlu F, Bilgiç V. Analysis of the Complexity Measures in the EEG of Schizophrenia Patients. International Journal of Neural Systems. Published online February 21, 2016. doi:10.1142/S0129065716500088

82. Karanikolaou M, Limanowski J, Northoff G. Does temporal irregularity drive prediction failure in schizophrenia? temporal modelling of ERPs. Schizophr. 2022;8(1):23. doi:10.1038/s41537-022-00239-7

83. Poza J, Hornero R, Abásolo D, Fernández A, Escudero J. Analysis of spontaneous MEG activity in patients with Alzheimer’s disease using spectral entropies. Annu Int Conf IEEE Eng Med Biol Soc. 2007;2007:6180–6183. doi:10.1109/IEMBS.2007.4353766

84. Gómez C, Hornero R. Entropy and Complexity Analyses in Alzheimer’s Disease: An MEG Study. Open Biomed Eng J. 2010;4:223–235. doi:10.2174/1874120701004010223

85. Ferdinando H, Moradi S, Korhonen V, Helakari H, Kiviniemi V, Myllylä T. Spectral entropy provides separation between Alzheimer’s disease patients and controls: a study of fNIRS. Eur Phys J Spec Top. 2023;232(5):655–662. doi:10.1140/epjs/s11734-022-00753-w

86. McGrath JJ, Lim CCW, Plana-Ripoll O, et al. Comorbidity within mental disorders: a comprehensive analysis based on 145 990 survey respondents from 27 countries. Epidemiol Psychiatr Sci. 2020;29:e153. doi:10.1017/S2045796020000633

87. Hu L, Chen H, Su W, et al. Aberrant static and dynamic functional connectivity of the executive control network in lung cancer patients after chemotherapy: a longitudinal fMRI study. Brain Imaging and Behavior. 2020;14(3):927–940. doi:10.1007/s11682-020-00287-6

88. Gorgolewski KJ, Durnez J, Poldrack RA. Preprocessed Consortium for Neuropsychiatric Phenomics dataset. F1000Res. 2017;6:1262. doi:10.12688/f1000research.11964.2

