## Supplementary Information for "A mega-analysis of low frequency resting-state measures in mood and psychosis-spectrum disorders"

**Supplementary Methods**

**Additional Dataset Information.** Participants in all four datasets underwent resting-state functional magnetic resonance imaging (rsfMRI). The COBRE^1^ dataset participants were recruited from the University of New Mexico Psychiatric Center and the Raymond G. Murphy Veterans Affairs Medical Center. This sample included 58 healthy controls (HCs), and 41 patients diagnosed with schizophrenia or schizoaffective disorder between 18 and 64 years of age. The HCP-EP^2^ quality-controlled dataset consisted of 112 early psychosis (EP) patients (within the first five years of the first presentation of psychosis symptoms) and 57 HCs aged 16-35 from four different recruitment sites: Beth Israel Deaconess Medical Center, McLean Hospital, and Massachusetts General Hospital, and Indiana University. The sample also includes affective and non-affective psychosis patients diagnosed according to the DSM-5^3^. Affective patients had a mood disorder with psychotic features (e.g., major depression with psychosis or bipolar disorder with psychosis)^2^. The SRPBS^4^ dataset is a transdiagnostic population of patients with mood and psychosis symptoms of varying severities. The final sample included HCs (n=303), individuals diagnosed with schizophrenia spectrum disorder (SSD, n=124), and individuals diagnosed with a mood spectrum disorder (n=182; bipolar with unknown type=46, dysthymia=2, and major depressive disorder=134) aged 16-80 from 7 different recruitment sites in Japan: Center of Innovation at Hiroshima University (COI), Hiroshima Kajikawa Hospital (HKH), Hiroshima Rehabilitation Center (HRC), Hiroshima University Hospital (HUH), Kyoto University (KUT), Showa University (SWA), and University of Tokyo Hospital (UTO). The CNP^5^ quality-controlled sample has 117 HCs, 41 schizophrenia, and 46 bipolar disorder (type unknown) participants from the local community aged 21-50. The bipolar samples in the SRPBS and UCLA datasets may include bipolar I, in which affected individuals may experience psychotic symptoms during mania. Given that these subjects were diagnosed using the DSM-5, they are classified as mood disorders.

**Additional Quality Control Information.** 1332 participants initially survived standard quality control. However, not all these subjects had full brain coverage on their fMRI data. 303 participants had missing or insufficient node and network coverage, but 288 of them had at least 75 percent coverage of member nodes of a given network. To maximize data utility, only the subjects with 100 percent of member nodes with at least 75 percent coverage for each network (e.g., all subjects with complete data for the medial frontal network would undergo analyses relevant to the medial frontal network) were used in subsequent analysis.

**Additional Site Harmonization Information**. Some networks had sites with only one participant, which meant that variance estimates could not be reliably estimated using the default *neuroCombat* algorithm. In these cases, the data were harmonized using the mean-only algorithm. For all other sites, harmonization controlling for mean and variance were used. We decided against excluding these subjects to maximize sample size as much as possible and to also reflect real-world conditions where only one quality-controlled sample is available for analysis.

**Analyses without influential data points.** Additionally, we identified influential participants using an internal R Studio function called *boxplot.stats*. Analyses were repeated without these points and reveal similar trends as the main analyses (Figure S4, Tables S8 and S11).

**Supplementary Results**

**ALFF and lfSE have a moderately inverse relationship.** ALFF and lfSE are moderately negatively associated (Tables S16). Consistent with this observation, at almost every level of analysis, ALFF and lfSE had opposite significance effect directions as well.

**Supplementary Figures**

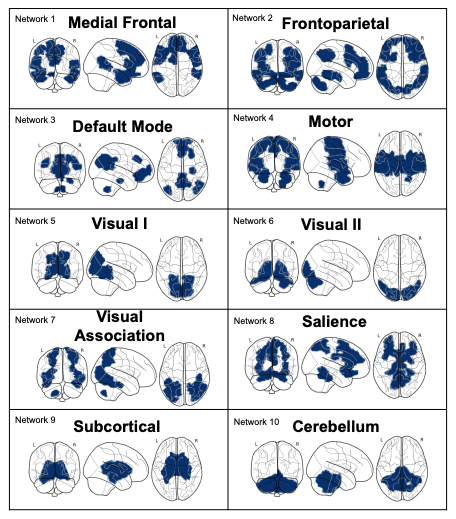

**Figure S1**. The ten canonical functional networks of the Shen-268 atlas.

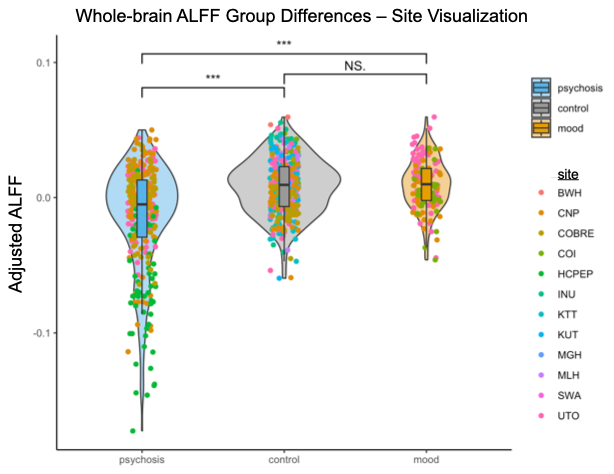
**Figure S2.** *ALFF Group Differences at Whole-brain Level Site Visualization.* Each dot represents a participant, and the color of the dot represents the site their data was collected from.

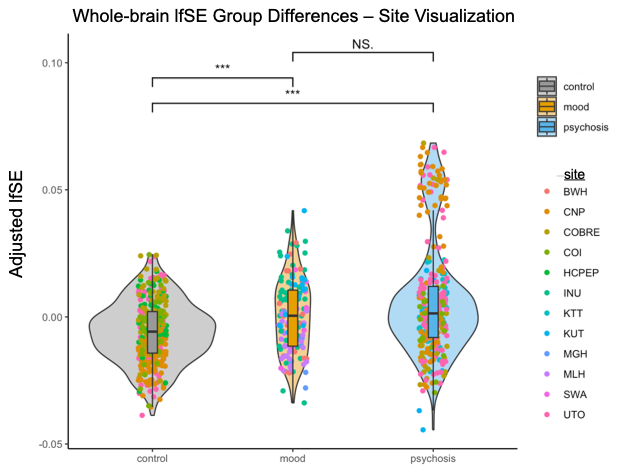

**Figure S3.** *lfSE Group Differences at Whole-brain Level Site Visualization.* Each dot represents a participant, and the color of the dot represents the site their data was collected from.

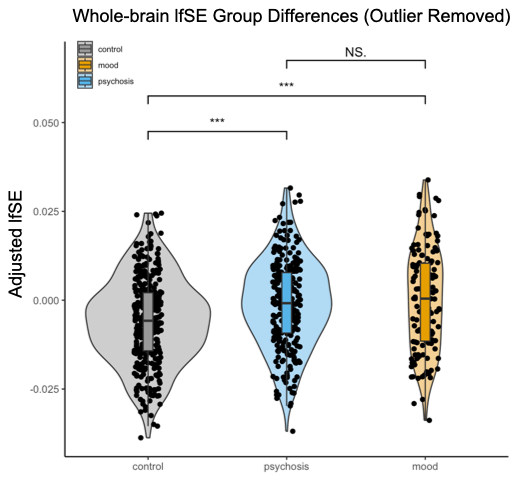
**Figure S4.** *lfSE Group Differences at Whole-brain Level with High Influence Participants Removed.* Violin plots of the results for ALFF group differences with high influence participant data removed Dots on the violin plots represent the subjects, colors represent the groups (controls=gray, mood=orange, psychosis=light blue). N.S.=not significant and ***= p<0.001. Controls still had significantly lower lfSE than mood and psychosis spectrum disorders. However, mood disorders have a higher average lfSE compared to psychosis disorders (though not significant), as compared to group differences with high influence participant data.

**Supplementary Tables**

| Demographic Variable | | Controls | Mood | Psychosis | Test Statistic |
| --- | --- | --- | --- | --- | --- |
| Sample Size | | 535 | 228 | 318 | N/A |
| Age (years) | | 39.89 ± 15.3 | 38.31 ± 12.56 | 29.8 ± 13.21 | W=75.48, df=2, p<0.001 |
| Motion Frame Displacement (mm) | | 0.083 ± 0.046 | 0.087 ± 0.045 | 0.075 ± 0.037 | W= 8.599, df=2, p=0.014 |
| Dataset sample contributions | | COBRE = 58  SRPBS = 303  HCP-EP = 57  UCLA = 117 | SRPBS = 182  UCLA = 46 | COBRE = 41  SRPBS = 124  HCP-EP = 112 UCLA = 41 | N/A |
| Self-reported sex (n=1081, F/M) | | 272/263 | 79/149 | 135/183 | χ²=18.081, df = 2, p<0.001 |
| BDI-II (n=62) | | 6.83 ± 6.39  (n=23) | 22.13 ± 12.17  (n=39) | N/A | F=3.31; df=1, p<0.01 |
| PANSS* (n=230) | Positive | N/A | N/A | 10.96 ± 3.81 | N/A |
|  | Negative | N/A | N/A | 13.98 ± 5.5 | N/A |
|  | General Psychopathology | N/A | N/A | 24.63 ± 5.34 | N/A |
|  | Total | N/A | N/A | 49.57 ± 10.93 | N/A |

**Table S1: Descriptive Statistics of Participant Demographics by Clinical Group.** Descriptive statistics of key demographic variables from the open-source datasets used in this study, including sex, motion, age, motion, and symptom score summaries. F=female; M=male. N/A = not applicable. PANSS=Positive and Negative Syndrome Scale, BDI-II =Beck's Depression Inventory. n=1081.

| **Dataset** | **Site** | **Scanner Type** | **Protocol** | **TR (sec)** | **# of frames** |
| --- | --- | --- | --- | --- | --- |
| COBRE | The University of New Mexico Psychiatric Center | Siemens 3T Prisma MRI System | Single-shot full k-space echo-planar imaging (with ramp sampling correction using the intercomissural line) | 2 | 150 |
|  | Raymond G. Murphy Veterans Affairs Medical Center | Siemens 3T Prisma MRI System | Single-shot full k-space echo-planar imaging (with ramp sampling correction using the intercomissural line) | 2 | 150 |
| HCP-EP | Brigham and Women's Hospital | Siemens MAGNETOM Prisma 3T | Multiband sequence and 32 channel head coils | 0.8 | 420 |
|  | Indiana University | Siemens MAGNETOM Prisma 3T | Multiband sequence and 32 channel head coils | 0.8 | 420 |
|  | Massachusetts General Hospital | Siemens MAGNETOM Prisma 3T | Multiband sequence and 64 channel head coils | 0.8 | 420 |
|  | McLean University | Siemens MAGNETOM Prisma 3T | Multiband sequence and 64 channel head coils | 0.8 | 420 |
| SRPBS | Hiroshima COI | Simens Verio.Dot 3T | Siemens ep2d_bold sequence and 12 channel headcoils | 2.5 | 240 |
|  | Kyoto University | Siemens TimTrio 3T | Siemens ep2d_bold sequence and 32 channel head coils | 2.5 | 240 |
|  | Hiroshima Kajikawa Hospital | Simens Spectra 3T | Siemens ep2d_bold sequence and 12 channel head coils | 2.7 | 107 |
|  | Hiroshima Rehabilitation Center | GE Signa HDxt 3T | Multiband GE‑EPI BOLD sequence and 8 channel head coils | 2 | 143 |
|  | Hiroshima University Hospital | GE Signa HDxt 3T | Multiband GE‑EPI BOLD sequence, and 8 channel head coils | 2 | 143 |
|  | Showa University | Siemens Verio 3T | Siemens ep2d_bold sequence and 12 channel head coils | 2.5 | 244 |
|  | University of Tokyo Hospital | General Electric MR750W 3T | Siemens ep2d_bold sequence equivalent and 24 channel head coils | 2.5 | 240 |
| CNP | University of California Los Angeles Semel Institute | Siemens TimTrio 3T | Siemens ep2d_bold sequence and 12 channel head coils | 2 | 152 |

**Table S2***. Site-Specific Scanner Information***.** Imaging protocols including acquisition parameters and scanner characteristics are reported for each site represented in the study. COBRE= Center for Biomedical Research Excellence, Human Connectome Project Early Psychosis =HCP-EP, Strategic Research Program for Brain Sciences =SRPBS, Consortium for Neuropsychiatric Phenomics=CNP.

| **Scale** | Total | Control | Mood | Psychosis |
| --- | --- | --- | --- | --- |
| Whole-brain | 823 | 366 | 139 (BPD=46) | 318 (AP=28, EP=112, NAP=79) |
| Medial Frontal | 846 | 381 | 147 (BPD=46) | 318 (AP=28, EP=112, NAP=79) |
| Frontoparietal | 852 | 383 | 150 (BPD=46) | 318 (AP=28, EP=112, NAP=79) |
| Default Mode | 882 | 404 | 160 (BPD=46) | 318 (AP=28, EP=112, NAP=79) |
| Motor | 881 | 401 | 160 (BPD=46) | 318 (AP=28, EP=112, NAP=79) |
| Visual I | 1081 | 535 | 224 (BPD=46, DYS=2) | 318 (AP=28, EP=112, NAP=79) |
| Visual II | 953 | 440 | 194 (BPD=46, DYS=1) | 318 (AP=28, EP=112, NAP=79) |
| Visual Association | 906 | 412 | 175 (BPD=46, DYS=1) | 318 (AP=28, EP=112, NAP=79) |
| Salience | 936 | 415 | 174 (BPD=46, DYS=2) | 318 (AP=28, EP=112, NAP=79) |
| Subcortical | 906 | 412 | 175 (BPD=46, DYS=1) | 318 (AP=28, EP=112, NAP=79) |
| Cerebellar | 823 | 366 | 139 (BPD=46) | 318 (AP=28, EP=112, NAP=79) |

**Table S3:** *Sample distributions by network and clinical group***.** Major depressive disorder comprises most of the mood sample. The other diagnostic categories bipolar I (BPD) and dysthymia (DYS) sample sizes are listed in parentheses. The psychosis sample is a mix of psychosis stages, as well as a mix of affective and non-affective subtypes. Known early stage (EP) samples and psychosis subtypes (NAP=non-affective psychosis. AP=affective psychosis) are also listed in paratheses.

| **Node Coverage Type** | **Num Subjects** | **Proportion of full dataset** |
| --- | --- | --- |
| Full | 823 | 0.76 |
| Partial (at least 75% Whole-brain coverage) | 288 | 0.24 |

**Table S4***. Node Coverage Information***.** Number of subjects with partial or full node coverage.

| **Race** | **HCP-EP (Patients/Controls)** | **SRPBS (Patients/Controls)** | **UCLA**  **(BP/SCHZ/Controls)** |
| --- | --- | --- | --- |
| American Indian/Alaska Native | 1/0 | N/A | 4/9/23 |
| Asian | 7/8 | 123/99 | 0/0/2 |
| Black or African American | 40/5 | N/A | 1/2/1 |
| Multiracial | 1/1 | N/A | 7/1/0 |
| Unknown/Not Reported | 3/2 | N/A | 0 |
| White | 55/41 | N/A | 34/28/91 |

### **Table S5:** *Subject Race Distribution*. Available information on racial make-up of study’s sample. HCP-EP = Human Connectome Project Early Psychosis, SRPBS = Strategic Research Program for Brain Sciences, UCLA= University of California Los Angeles. We did not have ethnic or racial information for the Center of Biomedical Research Excellence (COBRE) dataset. BP=Bipolar, SCHZ=Schizophrenia.

| **Medication Status** | **HCP-EP (Current)** | **UCLA (M/P)** | **COBRE** | **SRPBS**** |
| --- | --- | --- | --- | --- |
| Sample with Medication | 86 | 37/37 | 40 | 91 |
| Proportion of Sample with Medication Exposure | 0.77 | 0.36 | 0.98 | 0.24 |
| Anti-psychotic (atypical/typical) | 81 (N/A) | 14 (10/4) | 39 (37/2) | 85 |
| Anti-depressants/mood stabilizers | N/A | 20 | N/A | 29 |
| Other | N/A | 30 | 1 | 66 |

### **Table S6:** *Medication Exposure Information*. Available medication information on study sample. HCP-EP = Human Connectome Project Early Psychosis, the Center of Biomedical Research Excellence (COBRE) dataset, and UCLA = University of California Los Angeles. **We did not have complete medication information for the Strategic Research Program for Brain Sciences (SRPBS) dataset. Thus, only the KTT/KUT sites information is displayed. Current=currently on medication during study. M=mood, P=psychosis.

| **Measure** | **Group** | **Mean ± Standard Deviation** |
| --- | --- | --- |
| Amplitude of low-frequency fluctuation (ALFF) | Control | 0.85 ± 0.03 |
|  | Mood | 0.84 ± 0.03 |
|  | Psychosis | 0.83 ± 0.05 |
| Low-frequency spectral entropy (lfSE) | Control | 0.61 ± 0.06 |
|  | Mood | 0.65 ± 0.04 |
|  | Psychosis | 0.61 ± 0.08 |

**Table S7***.* *Empirical ranges of ALFF and lfSE by clinical group*. Value distributions of non-covariate regressed ALFF and lfSE by clinical group.

|  |  | Clinical Groups (median +/- std) | | | Wilcoxon Rank Pairwise p | | |
| --- | --- | --- | --- | --- | --- | --- | --- |
| **Scale** | n | C | M | P | C vs M | C vs P | M vs P |
| Whole-brain | N = 799 (control outliers=3, mood outliers=3, psychosis outliers=18) | 0.00894 ± 0.0201 | 0.0102 ± 0.0172 | -0.00772 ± 0.0299 | p = 0.548  q = 0.548 | **p = 2.98e-12**  **q = 8.95e-12** | **p = 2.49e-09 q = 3.73e-09** |
| Medial Frontal | N = 820 (control outliers=3, mood outliers=1, psychosis outliers=22) | 0.00861 ± 0.0239 | 0.0115 ± 0.0237 | -0.00695 ± 0.0309 | p = 0.223  q = 0.223 | **p = 6.18e-10**  **q = 1.85e-09** | **p = 7.06e-09**  **q = 1.06e-08** |
| Frontoparietal | N = 835 (mood outliers=4, psychosis outliers=13) | 0.00749 ± 0.0221 | 0.0104 ± 0.019 | -0.00844 ± 0.0328 | p = 0.119  q = 0.119 | **p = 8.93e-1**  **q = 1.86e-09** | **p = 1.24e-09 q = 1.86e-09** |
| Default Mode | N = 846 (control outliers=6, mood outliers=1 psychosis outliers=29) | 0.00849 ± 0.0239 | 0.0112 ± 0.0247 | -0.00354 ± 0.0287 | p = 0.137  q = 0.137 | **p = 2.34e-07**  **q = 3.5e-07** | **p = 1.57e-07**  **q = 3.5e-07** |
| Motor | N = 846 (control outliers=8, mood outliers=3, psychosis outliers=24) | 0.009 ± 0.0222 | 0.0112 ± 0.022 | -0.0068 ± 0.0304 | p = 0.348  q = 0.348 | **p = 3.36e-11 q = 1.01e-10** | **p = 2.4e-09**  **q = 3.61e-09** |
| Visual I | N = 1054 (control outliers=16, mood outliers=3, psychosis outliers=8) | 0.00635 ± 0.0247 | 0.00913 ± 0.0276 | -0.00956 ± 0.0389 | p = 0.13  q = 0.13 | **p = 2.33e-07**  **q = 3.5e-07** | **p = 1.84e-07**  **q = 3.5e-07** |
| Visual II | N = 937 (control outliers=9, mood outliers=2, psychosis outliers=5) | 0.00819 ± 0.0269 | 0.00837 ± 0.0299 | -0.011 ± 0.0421 | p = 0.878  q = 0.878 | **p = 5.44e-10 q = 1.63e-09** | **p = 4.55e-07**  **q = 6.82e-07** |
| Visual Association | N = 881 (control outliers=3, mood outliers=5, psychosis outliers=17) | 0.00812 ± 0.0243 | 0.0119 ± 0.0255 | -0.00917 ± 0.0346 | p = 0.0993  q = 0.0993 | **p = 1.07e-10**  **q = 3.2e-10** | **p = 3.46e-10**  **q = 5.19e-10** |
| Salience | N = 897 (control outliers=4, mood outliers=4, psychosis outliers=31) | 0.00683 ± 0.0226 | 0.00934 ± 0.0227 | -0.00425 ± 0.026 | p = 0.191  q = 0.191 | **p = 2.01e-07**  **q = 3.01e-07** | **p = 3.47e-08**  **q = 1.04e-07** |
| Subcortical | N = 878 (control outliers=7, mood outliers=5, psychosis outliers=16) | 0.00868 ± 0.0231 | 0.00959 ± 0.023 | -0.00976 ± 0.0338 | p = 0.732  q = 0.732 | **p = 1.39e-12 q = 4.18e-12** | **p = 3.02e-09 q = 4.52e-09** |
| Cerebellar | N = 811 (control outliers=1, mood outliers=2, psychosis outliers=9) | 0.0114 ± 0.0315 | 0.0122 ± 0.0266 | -0.0125 ± 0.0458 | p = 0.836  q = 0.836 | **p = 1.22e-11**  **q = 3.65e-11** | **p = 1.16e-07 q = 1.74e-07** |

**Table S8.** *Effect of High Influence Group Member Removal on Whole-brain ALFF Group Differences.* High influence participants, calculated as outliers using an internal R Studio function, were removed to see if the observed group-level trends changed. C=controls, M=mood, P=psychosis.

|  |  | Clinical Groups (median +/- std) | | | Wilcoxon Rank Pairwise p | | |
| --- | --- | --- | --- | --- | --- | --- | --- |
| **Scale** | n | C | M | P | C vs M | C vs P | M vs P |
| Whole-brain | 795 | 0.008 ± 0.021 | 0.01 ± 0.019 | -0.015 ± 0.04 | p = 0.543  q = 0.543 | **p = 3.87e-15**  **q = 1.16e-14** | **p = 4.15e-11**  **q = 6.23e-11** |
| Medial Frontal | 818 | 0.008 ± 0.025 | 0.011 ± 0.025 | -0.016 ± 0.042 | p = 0.221  q = 0.221 | **p = 2.04e-13**  **q = 6.13e-13** | **p = 3.64e-11**  **q = 5.45e-11** |
| Frontoparietal | 825 | 0.007 ± 0.022 | 0.009 ± 0.022 | -0.015 ± 0.042 | p = 0.195  q = 0.195 | **p = 4.21e-12**  **q = 1.26e-11** | **p = 1.87e-10**  **q = 2.8e-10** |
| Default Mode | 854 | 0.008 ± 0.026 | 0.011 ± 0.025 | -0.016 ± 0.047 | p = 0.126  q = 0.126 | **p = 1.34e-11**  **q = 4.02e-11** | **p = 1.23e-10**  **q = 1.85e-10** |
| Motor | 853 | 0.008 ± 0.025 | 0.011 ± 0.024 | -0.017 ± 0.043 | p = 0.282  q = 0.282 | **p = 2.28e-15**  **q = 6.85e-15** | **p = 1.78e-12**  **q = 2.67e-12** |
| Visual I | 1054 | 0.004 ± 0.028 | 0.008 ± 0.029 | -0.014 ± 0.044 | p = 0.0807 q = 0.0807 | **p = 5.84e-07**  **q = 8.75e-07** | **p = 1.01e-07**  **q = 3.04e-07** |
| Visual II | 926 | 0.006 ± 0.03 | 0.007 ± 0.031 | -0.014 ± 0.046 | p = 0.722  q = 0.722 | **p = 1.5e-09**  **q = 4.49e-09** | **p = 2.35e-07**  **q = 3.52e-07** |
| Visual Association | 878 | 0.008 ± 0.025 | 0.01 ± 0.029 | -0.017 ± 0.046 | p = 0.184  q = 0.184 | **p = 6.24e-14**  **q = 1.87e-13** | **p = 1.62e-11**  **q = 2.43e-11** |
| Salience | 818 | 0.006 ± 0.024 | 0.009 ± 0.025 | -0.016 ± 0.041 | p = 0.171  q = 0.171 | **p = 3.24e-13**  **q = 9.71e-13** | **p = 1.51e-12**  **q = 2.26e-12** |
| Subcortical | 825 | 0.007 ± 0.025 | 0.011 ± 0.028 | -0.017 ± 0.043 | p = 0.328  q = 0.328 | **p = 1.2e-14**  **q = 3.61e-14** | **p = 4.07e-12**  **q = 6.11e-12** |
| Cerebellar | 854 | 0.011 ± 0.032 | 0.011 ± 0.029 | -0.019 ± 0.055 | p = 0.685  q = 0.685 | **p = 3.55e-13 q = 1.06e-12** | **p = 3.79e-08**  **q = 5.69e-08** |

**Table S9.** *Effect of Affective Psychosis Removal on Whole-brain ALFF Group Differences.* A subsample of the psychosis sample had a known affective psychosis disorder (n=28). These subjects were removed to test if they influenced group differences. Significant effects remained consistent with analyses including affective psychosis conditions. C=controls, M=mood, P=psychosis.

| **Scale** | n | Clinical Groups (median +/- std) | | | Wilcoxon Rank Pairwise p | | |
| --- | --- | --- | --- | --- | --- | --- | --- |
|  |  | C | M | P | C vs M | C vs P | M vs P |
| Whole-brain | 795 | -0.006 ± 0.012 | 0 ± 0.015 | 0.007 ± 0.023 | **p = 2.14e-05**  **q = 3.21e-05** | **p = 1.38e-13**  **q = 4.14e-13** | p = 0.0889  q = 0.0889 |
| Medial Frontal | 818 | -0.006 ± 0.014 | -0.002 ± 0.015 | 0.009 ± 0.028 | **p = 0.00523**  **q = 0.00523** | **p = 1.83e-13**  **q = 5.49e-13** | **p = 0.00334**  **q = 0.00501** |
| Frontoparietal | 825 | -0.005 ± 0.014 | -0.001 ± 0.018 | 0.007 ± 0.025 | **p = 0.00145**  **q = 0.00218** | **p = 1.54e-10 q = 4.61e-10** | **p = 0.025**  **q = 0.025** |
| Default Mode | 854 | -0.004 ± 0.016 | -0.004 ± 0.019 | 0.007 ± 0.031 | p = 0.218  q = 0.218 | **p = 4.53e-07 q = 1.36e-06** | **p = 0.0036**  **q = 0.0054** |
| Motor | 853 | -0.006 ± 0.014 | -0.003 ± 0.019 | 0.009 ± 0.028 | **p = 0.00583**  **q = 0.00799** | **p = 1.43e-10 q = 4.29e-10** | **p = 0.00799**  **q = 0.00799** |
| Visual I | 1054 | -0.002 ± 0.017 | -0.009 ± 0.03 | 0.01 ± 0.026 | p = 0.0668  q = 0.0668 | **p = 7.97e-10 q = 1.19e-09** | **p = 6.69e-10 q = 1.19e-09** |
| Visual II | 926 | -0.003 ± 0.018 | -0.004 ± 0.027 | 0.008 ± 0.028 | p = 0.177  q = 0.177 | **p = 2e-08**  **q = 6.01e-08** | **p = 0.000654 q = 0.000982** |
| Visual Association | 878 | -0.005 ± 0.016 | -0.006 ± 0.027 | 0.011 ± 0.029 | p = 0.151  q = 0.151 | **p = 4.3e-13 q = 1.29e-12** | **p = 2.64e-05 q = 3.96e-05** |
| Salience | 818 | -0.004 ± 0.013 | -0.004 ± 0.021 | 0.009 ± 0.027 | p = 0.0825  q = 0.0825 | **p = 1.83e-10 q = 5.49e-10** | **p = 0.000205 q = 0.000307** |
| Subcortical | 825 | -0.005 ± 0.013 | -0.005 ± 0.026 | 0.01 ± 0.027 | **p = 0.00239**  **q = 0.00358** | **p = 2.43e-13 q = 7.29e-13** | **p = 0.00805**  **q = 0.00805** |
| Cerebellar | 854 | -0.007 ± 0.019 | 0.002 ± 0.019 | 0.007 ± 0.035 | **p = 7.93e-06 q = 1.19e-05** | **p = 6.14e-09 q = 1.84e-08** | p = 0.41  q = 0.41 |

**Table S10.** *Effect of Affective Psychosis Removal on Whole-brain lfSE Group Differences.* A subsample of the psychosis sample had a known affective psychosis disorder (n=28). These subjects were removed to test if they influenced group differences. Significant effects remained consistent with analyses including affective psychosis conditions.

| **Scale** | n | Clinical Groups (median +/- std) | | | Wilcoxon Rank Pairwise p | | |
| --- | --- | --- | --- | --- | --- | --- | --- |
|  |  | C | M | P | C vs M | C vs P | M vs P |
| Whole-brain | N = 780 (psychosis outliers=43) | -0.00595 ± 0.0121 | 0.000456 ± 0.0147 | -0.00061 ± 0.0133 | **p = 2.14e-05 q = 3.21e-05** | **p = 6.26e-07 q = 1.88e-06** | p = 0.476  q = 0.476 |
| Medial Frontal | N = 798 (control outliers=3, psychosis outliers=45) | -0.00575 ± 0.0134 | -0.00221 ± 0.0155 | -0.000527 ± 0.0143 | **p = 0.00758**  **q = 0.0114** | **p = 2.62e-06**  **q = 7.86e-06** | p = 0.527  q = 0.527 |
| Frontoparietal | N = 822 (control outliers=3, mood outliers=1, psychosis outliers=26) | -0.00489 ± 0.0135 | -0.000131 ± 0.0158 | 0.00285 ± 0.0191 | **p = 0.00139 q = 0.00208** | **p = 3.02e-07**  **q = 9.05e-07** | p = 0.318  q = 0.318 |
| Default Mode | N = 832 (control outliers=3, mood outliers=4, psychosis outliers=43) | -0.0039 ± 0.0155 | -0.00224 ± 0.015 | 0.00143 ± 0.0171 | p = 0.175  q = 0.175 | **p = 0.000216**  **q = 0.000649** | p = 0.101  q = 0.152 |
| Motor | N = 828 (control outliers=3, mood outliers=2, psychosis outliers=48) | -0.00515 ± 0.0137 | -0.00133 ± 0.015 | -0.000665 ± 0.0133 | **p = 0.00385**  **q = 0.00577** | **p = 7.13e-05**  **q =0.000214** | p = 0.844  q = 0.844 |
| Visual I | N = 1049 (control outliers=1, mood outliers=19, psychosis outliers=12) | -0.00145 ± 0.0173 | -0.00221 ± 0.0192 | 0.00755 ± 0.0229 | p = 0.961  q = 0.961 | **p = 5.93e-08 q = 1.78e-07** | **p = 2.09e-05**  **q = 3.14e-05** |
| **Visual II** | N = 934 (control outliers=2, mood outliers=8, psychosis outliers=9) | **-0.00307 ± 0.0176** | **-0.000199 ± 0.0187** | **0.00719 ± 0.0246** | **p = 0.0314**  **q = 0.0314** | **p = 1.39e-08**  **q = 4.18e-08** | **p = 0.00659**  **q = 0.00988** |
| Visual Association | N = 850 (control outliers=10, mood outliers=8, psychosis outliers=38) | -0.00451 ± 0.0148 | -0.00158 ± 0.0184 | 0.00236 ± 0.0168 | **p = 0.0311**  **q = 0.0467** | **p = 3.45e-07 q = 1.03e-06** | p = 0.105  q = 0.105 |
| Salience | N = 878 (control outliers=2, mood outliers=8, psychosis outliers=48) | -0.00427 ± 0.0129 | -0.000808 ± 0.0125 | -0.000776 ± 0.0118 | **p = 0.0101**  **q = 0.0152** | **p = 0.000894**  **q = 0.00268** | p = 0.771  q = 0.771 |
| Subcortical | N = 847 (control outliers=3, mood outliers=8, psychosis outliers=48) | -0.00471 ± 0.0123 | -0.000455 ± 0.015 | -0.000238 ± 0.00975 | **p = 8.95e-05 q = 0.000134** | **p = 1.33e-05**  **q = 4e-05** | p = 0.297  q = 0.297 |
| Cerebellar | N = 776 (control outliers=5, mood outliers=3, psychosis outliers=39) | -0.00639 ± 0.0175 | 0.00252 ± 0.0175 | 0.000491 ± 0.0218 | **p = 5.04e-06 q = 1.51e-05** | **p = 6.53e-05**  **q = 9.8e-05** | p = 0.285  q = 0.285 |

**Table S11.** *Effect of High-Influence Group Member Removal on Whole-brain lfSE Group Differences.* High influence participants, calculated as outliers using an internal R Studio function, were removed to see if the observed group-level trends changed. Psychosis and mood lfSE did reverse (see Table 3). Controls still had significantly lower whole-brain lfSE than mood and psychosis disorders. Visual II group differences between all three groups were significant once outliers were removed. C=controls, M=mood, P=psychosis.

| **Scale** | **Sample Size** | **Spearman’s ρ** |
| --- | --- | --- |
| Whole-brain with only Full Nodes | 32 | 0.20 (p=0.2841*) |
| Whole-brain with Available Nodes | 62 | **0.36 (p=0.0046*)** |
| Medial Frontal | 48 | 0.05 (p=0.747*,0.7470) |
| Frontoparietal | 41 | 0.07 (p=0.6616*, 0.7470) |
| Default Mode | 58 | 0.11 (p=0.3926*, 0.7470) |
| Motor | 35 | 0.06 (p=0.7332*, 0.7470) |
| Visual I | 57 | 0.05 (p=0.7339*, 0.7470) |
| Visual II | 56 | -0.06 (p=0.6591*, 0.7470) |
| Visual Association | 44 | -0.07 (p=0.6619*, 0.7470) |
| Salience | 49 | 0.15 (p=0.3047*, 0.7470) |
| Subcortical | 56 | 0.09 (p=0.4894*, 0.7470) |
| Cerebellar | 32 | -0.11 (p=0.5379*, 0.7470) |

**Table S12***. Spearman’s rho associations between BDI-II scores and across scales of analysis.* *=uncorrected p-values. Participants were either matched healthy controls or had a mood disorder. All participants were from one site in the SRPBS dataset. “Whole-brain with only Full Nodes” were individuals with full data for all 268 nodes. “Whole-brain with Available Nodes” was calculated using the available full node data for all BDI-II score individuals.

| **Scale** | **Spearman’s ρ** |
| --- | --- |
| Whole-brain | -0.06 (p=0.387*) |
| Medial Frontal | -0.05 (p=0.4900*, q=0.832) |
| Frontoparietal | -0.10 (p=0.1340*, q=0.0.472) |
| Default Mode | -0.01 (p=0.8720*, q=0.872) |
| Motor | -0.04 (p=0.5350*, q=0.8320) |
| Visual I | -0.10 (p=0.1430*, q=0.4720) |
| Visual II | -0.09 (p=0.1890*, q=0.4720) |
| Visual Association | -0.13 (p=0.0560*, q=0.4720) |
| Salience | -0.04 (p=0.5820*, q=0.832) |
| Subcortical | -0.01 (p=0.8620*, q=0.872) |
| Cerebellar | 0.03 (p=0.6860*, q=0.857) |

**Table S13***.* *Spearman’s rho associations between positive PANSS scores and ALFF across scales of analysis.* *=uncorrected, q=FDR corrected p-value. n=230 (HCP-EP=107, SRPBS=123).

| Scale | Spearman’s ρ |
| --- | --- |
| Whole-brain | **-0.13 (p=0.0418*)** |
| Medial Frontal | -0.10 (p=0.1460*, q=0.1620) |
| Frontoparietal | -0.12 (p=0.0610*, q=0.1330) |
| Default Mode | -0.10 (p=0.1140*, q=0.1430) |
| Motor | -0.13 (p= 0.0430*, q=0.1330) |
| Visual I | -0.09 (p= 0.1790*, q=0.1790) |
| Visual II | -0.13 (p= 0.0420*, q=0.1330) |
| Visual Association | -0.11 (p= 0.0830*, q=0.1330) |
| Salience | -0.11 (p= 0.0930*, q=0.1330) |
| Subcortical | -0.11 (p=0.0850*, q=0.1330) |
| Cerebellar | -0.14 (p= 0.0290*, q=0.1330) |

**Table S14***.* *Spearman’s rho associations between positive PANSS scores and lfSE across scales of analysis*. *=uncorrected, q=FDR corrected p-value. n=230 (HCP-EP=107, SRPBS=123). Bolded table entries represent significant comparisons.

| Scale | Sample Size | Spearman’s ρ |
| --- | --- | --- |
| Whole-brain with only Full Nodes | 32 | **-0.45 (p=0.0103*)** |
| Whole-brain with Available Nodes | 62 | **-0.38 (p=0.00222*)** |
| Medial Frontal | 48 | -0.21 (p=0.1473*,0.2798) |
| Frontoparietal | 41 | 0.001 (p=0.9951*,0.9951) |
| Default Mode | 58 | -0.22 (p=0.1030*,0.2798) |
| Motor | 35 | 0.30 (p=0.08425*,0.2798) |
| Visual I | 57 | -0.18 (p=0.19815*,0.2831) |
| Visual II | 56 | -0.21 (p=0. 0.1337*,0.2798) |
| Visual Association | 44 | -0.09 (p=0.5889*,0.6543) |
| Salience | 49 | -0.20 (p=0.1679*,0.2798) |
| Subcortical | 56 | -0.22 (p=0.1156*,0.2798) |
| Cerebellar | 32 | 0.16 (p=0.3968*,0.4961) |

**Table S15***.* *Spearman’s rho associations between BDI-II scores and lfSE across scales of analysis*. *=uncorrected p-values. Participants were either matched healthy controls or had a mood disorder. All participants were from one site in the SRPBS dataset. “Whole-brain with only Full Nodes” were individuals with full data for all 268 nodes. “Whole-brain with Available Nodes” was calculated using the available full node data for all BDI-II score individuals.

| Scale | All | Control | Mood | Psychosis |
| --- | --- | --- | --- | --- |
| Whole-brain | **-0.28 (p=2.00E-16)** | **-0.21 (p=5.122e-05)** | -0.087 (p=0.3081) | **-0.23 (p=3.027e-05)** |
| Medial Frontal | **-0.31 (p=6.43E-20)** | **-0.12 (p=0.01727)** | **-0.27 (p=0.001344)** | **-0.27 (p=1.412e-06)** |
| Frontoparietal | **-0.32 (p=0)** | **-0.18 (p=0.0004611)** | 0.12 (p= 0.1642) | **-0.15 (p=0.009207** |
| Default Mode | **-0.22 (p=6.29E-11)** | **-0.25 (p=** **1.07e-06)** | -0.15 (p=0.08818) | **-0.29 (p=1.232e-07)** |
| Motor | **-0.38 (p=0)** | **-0.24 (p=4.896e-06)** | **-0.26 (p=0.001762)** | **-0.22 (p=9.944e-05)** |
| Visual I | **-0.33 (p=0)** | **-0.30 (p=5.583e-09)** | **-0.22 (p=0.007928)** | **-0.33 (p=2.387e-09)** |
| Visual II | **-0.32 (p=0)** | **-0.25 (p=1.273e-06)** | **-0.35 (p=2.856e-05)** | **-0.32 (p=4.283e-09)** |
| Visual Association | **-0.33 (p=0)** | **-0.20 (p=8.111e-05)** | -0.07 (p=0.4275) | **-0.27 (p=1.419e-06)** |
| Salience | **-0.39 (p=0)** | **-0.18 (p=0.000774)** | **-0.45 (p= 2.844e-08)** | **-0.28 (p=6.955e-07)** |
| Subcortical | **-0.36 (p=0)** | **-0.10 (p=0.04755)** | 0.05 (p=0.5795) | **-0.26 (p=2.098e-06)** |
| Cerebellar | **-0.33 (p=0)** | 0.086 (p=0.1006) | 0.025 (p=0.7734) | **-0.16 (p=0.0.004731)** |

**Table S16***.* *Associations between ALFF and lfSE by scale of analysis and group*. ALFF and lfSE were correlated using Spearman’s rho using the average values of each scale of analysis per group (all together and independently). For example, control ALFF and control lfSE were correlated at each scale of analysis. The same were repeated for mood and psychosis disorders. p=p-value.

| Scale | Control vs Mood | Control vs Psychosis | Mood vs Psychosis |
| --- | --- | --- | --- |
| Whole-brain | Z=-1.253, p=0.105 | Z=0.273, p=0.392 | Z=1.432, p=0.076 |
| Medial Frontal | Z=1.596, p=0.055 | **Z=2.049, p=0.02** | Z=0, p=0.5 |
| Frontoparietal | **Z=-3.115, p=0.001** | Z=-0.405, p=0.343 | **Z=2.72, p=0.003** |
| Default Mode | Z=-1.108, p=0.134 | Z=0.573, p=0.283 | Z=1.509, p=0.066 |
| Motor | Z=-0.226, p=0.41 | Z=0.28, p=0.39 | Z=0.435, p=0.332 |
| Visual I | Z=-1.073, p=0.142 | Z=-0.469, p=0.32 | Z=-1.358, p=0.087 |
| Visual II | Z=1.269, p=0.102 | Z=1.031, p=0.151 | Z=-0.369, p=0.356 |
| Visual Association | Z=-1.459, p=0.072 | Z=0.989, p=0.161 | **Z=2.181, p=0.015** |
| Salience | **Z=3.328, p=0** | Z=1.412, p=0.079 | **Z=-2.074, p=0.019** |
| Subcortical | **Z=-1.655, p=0.049** | **Z=2.211, p=0.014** | **Z=3.335, p=0** |
| Cerebellar | Z=0.609, p=0.271 | **Z=3.215, p=0.001** | **Z=-1.817, p=0.035** |

**Table S17***.* ALFF-lfSE effect size comparisons. For each group pair, their ALFF-lfSE Spearman’s rho was compared, and a z-score was calculated. Significance was determined by p<0.05.

| Scale | n | Clinical Group | | | Wilcoxon Rank Pairwise p | | |
| --- | --- | --- | --- | --- | --- | --- | --- |
|  |  | Control | Mood | Psychosis | C vs M | C vs P | M vs P |
| Whole-brain | 823 | 0.008 ± 0.021 | 0.01 ± 0.019 | -0.014 ± 0.039 | p = 0.399  q = 0.399 | **p = 5.88e-15 q = 1.77e-14** | **p = 1.83e-11 q = 2.74e-11** |
| Medial Frontal | 846 | 0.008 ± 0.026 | 0.011 ± 0.026 | -0.015 ± 0.041 | p = 0.16  q = 0.16 | **p = 1.68e-13 q = 5.05e-13** | **p = 2.43e-11 q = 3.65e-11** |
| Frontoparietal | 852 | 0.008 ± 0.022 | 0.009 ± 0.023 | -0.014 ± 0.041 | p = 0.175  q = 0.175 | **p = 1.59e-12 q = 4.76e-12** | **p = 1.27e-10 q = 1.9e-10** |
| Default Mode | 882 | 0.008 ± 0.027 | 0.011 ± 0.026 | -0.015 ± 0.046 | p = 0.127  q = 0.127 | **p = 9.81e-12 q = 2.94e-11** | **p = 1.47e-10 q = 2.21e-10** |
| Motor | 881 | 0.008 ± 0.026 | 0.011 ± 0.025 | -0.015 ± 0.042 | p = 0.204  q = 0.204 | **p = 1.92e-14 q = 5.76e-14** | **p = 2.49e-12 q = 3.73e-12** |
| Visual I | 1081 | 0.005 ± 0.029 | 0.005 ± 0.029 | -0.012 ± 0.046 | p = 0.899  q = 0.899 | **p = 1.04e-05 q = 3.12e-05** | **p = 0.000194 q = 0.000291** |
| Visual II | 953 | 0.007 ± 0.031 | 0.007 ± 0.032 | -0.013 ± 0.046 | p = 0.953  q = 0.953 | **p = 1.56e-09 q = 4.69e-09** | **p = 1.92e-06 q = 2.88e-06** |
| Visual Association | 906 | 0.008 ± 0.026 | 0.008 ± 0.03 | -0.015 ± 0.045 | p = 0.66  q = 0.66 | **p = 2.4e-13 q = 7.19e-13** | **p = 3.09e-09 q = 4.63e-09** |
| Salience | 936 | 0.007 ± 0.025 | 0.009 ± 0.026 | -0.014 ± 0.04 | p = 0.3  q = 0.3 | **p = 6.89e-13 q = 2.07e-12** | **p = 5.91e-11**  **q = 8.87e-11** |
| Subcortical | 906 | 0.008 ± 0.027 | 0.008 ± 0.028 | -0.015 ± 0.042 | p = 0.888  q = 0.888 | **p = 1.24e-12 q = 3.73e-12** | **p = 1e-08**  **q = 1.51e-08** |
| Cerebellar | 823 | 0.011 ± 0.033 | 0.012 ± 0.029 | -0.017 ± 0.053 | p = 0.998  q = 0.998 | **p = 6.45e-13 q = 1.93e-12** | **p = 1.17e-08 q = 1.75e-08** |

**Table S18.** *Amplitude of low-frequency (ALFF, 0.01-0.045 Hz) value distributions by whole-brain, network and clinical group.* Reported values have had covariates linearly regressed and undergone site harmonization. n=sample size. The medial frontal, default mode, motor, visual association, salience, and subcortical networks were harmonized using the mean only algorithm.

| Scale | n | Clinical Group | | | Wilcoxon Rank Pairwise p | | |
| --- | --- | --- | --- | --- | --- | --- | --- |
|  |  | Control | Mood | Psychosis | C vs M | C vs P | M vs P |
| Whole-brain | 823 | 0.008 ± 0.021 | 0.01 ± 0.019 | -0.014 ± 0.039 | p = 0.399  q = 0.399 | **p = 5.88e-15 q = 1.77e-14** | **p = 1.83e-11 q = 2.74e-11** |
| Medial Frontal | 846 | 0.009 ± 0.089 | 0.012 ± 0.099 | -0.016 ± 0.046 | p = 0.0924  q = 0.0924 | **p = 9.12e-16 q = 2.73e-15** | **p = 6.7e-14**  **q = 1e-13** |
| Frontoparietal | 852 | 0.014 ± 0.054 | 0.009 ± 0.074 | -0.021 ± 0.073 | p = 0.725  q = 0.725 | **p = 3.12e-12 q = 9.36e-12** | **p = 3.3e-08**  **q = 4.95e-08** |
| Default Mode | 882 | 0.013 ± 0.115 | 0.002 ± 0.142 | -0.017 ± 0.058 | p = 0.555  q = 0.555 | **p = 1.49e-09 q = 4.47e-09** | **p = 7.23e-07 q = 1.08e-06** |
| Motor | 881 | 0.012 ± 0.111 | 0.005 ± 0.131 | -0.018 ± 0.055 | p = 0.575  q = 0.575 | **p = 7.33e-10 q = 2.2e-09** | **p = 2.06e-07 q = 3.09e-07** |
| Visual I | 1081 | 0.011 ± 0.128 | -0.035 ± 0.166 | 0.007 ± 0.143 | **p = 0.00743 q = 0.0129** | p = 0.712  q = 0.712 | **p = 0.00862**  **q = 0.0129** |
| Visual II | 953 | 0.017 ± 0.107 | -0.013 ± 0.145 | -0.015 ± 0.113 | p = 0.195  q = 0.195 | **p = 0.00189**  **q = 0.00566** | p = 0.174  q = 0.195 |
| Visual Association | 906 | 0.014 ± 0.122 | -0.028 ± 0.174 | -0.002 ± 0.068 | p = 0.214  q = 0.214 | **p = 2.59e-05 q = 7.78e-05** | p = 0.126  q = 0.189 |
| Salience | 936 | 0.017 ± 0.128 | -0.015 ± 0.171 | -0.014 ± 0.057 | p = 0.279  q = 0.279 | **p = 5.01e-05 q = 0.00015** | p = 0.0505  q = 0.0757 |
| Subcortical | 906 | 0.013 ± 0.111 | -0.026 ± 0.158 | -0.003 ± 0.063 | p = 0.149  q = 0.149 | **p = 1.57e-06 q = 4.7e-06** | p = 0.0709  q = 0.106 |
| Cerebellar | 823 | 0.011 ± 0.033 | 0.012 ± 0.029 | -0.017 ± 0.053 | p = 0.998  q = 0.998 | **p = 6.45e-13 q = 1.93e-12** | **p = 1.17e-08 q = 1.75e-08** |

**Table S19.** *Amplitude of low-frequency (ALFF, 0.035-0.090 Hz) value distributions by whole-brain, network and clinical group.* Reported values have had covariates linearly regressed and undergone site harmonization. n=sample size. The medial frontal, default mode, motor, visual association, salience, and subcortical networks were harmonized using the mean only algorithm.

|  |  | Clinical Groups (median +/- std) | | | Wilcoxon Rank Pairwise p | | |
| --- | --- | --- | --- | --- | --- | --- | --- |
| Scale | n | Controls | Mood | Psychosis | C vs M | C vs P | M vs P |
| Whole-brain | 823 | -0.005 ± 0.014 | -0.002 ± 0.018 | 0.007 ± 0.023 | p = 0.061  q = 0.061 | **p = 2.84e-12 q = 8.52e-12** | **p = 0.00107**  **q = 0.0016** |
| Medial Frontal | 846 | -0.006 ± 0.016 | -0.004 ± 0.019 | 0.009 ± 0.028 | p = 0.195  q = 0.195 | **p = 4.32e-12 q = 1.3e-11** | **p = 0.000219**  **q = 0.000329** |
| Frontoparietal | 853 | -0.005 ± 0.016 | -0.002 ± 0.02 | 0.007 ± 0.025 | p = 0.127  q = 0.127 | **p = 1.71e-10 q = 5.13e-10** | **p = 0.00107**  **q = 0.0016** |
| Default Mode | 882 | -0.005 ± 0.018 | -0.003 ± 0.02 | 0.008 ± 0.03 | p = 0.319  q = 0.319 | **p = 7.43e-09 q = 2.23e-08** | **p = 0.000434**  **q = 0.000651** |
| Motor | 881 | -0.005 ± 0.016 | -0.003 ± 0.021 | 0.008 ± 0.027 | p = 0.257  q = 0.257 | **p = 7.86e-11 q = 2.36e-10** | **p = 0.000271**  **q = 0.000407** |
| Visual I | 1082 | -0.001 ± 0.018 | -0.005 ± 0.022 | 0.006 ± 0.026 | **p = 0.0219**  **q = 0.0219** | **p = 0.000342 q = 0.000513** | **p = 4.81e-06**  **q = 1.44e-05** |
| Visual II | 954 | -0.004 ± 0.021 | -0.002 ± 0.024 | 0.007 ± 0.027 | p = 0.0868  q = 0.0868 | **p = 3.18e-08 q = 9.53e-08** | **p = 0.00481**  **q = 0.00721** |
| Visual Association | 906 | -0.005 ± 0.018 | -0.003 ± 0.022 | 0.008 ± 0.028 | p = 0.0676  q = 0.0676 | **p = 1.62e-09 q = 4.85e-09** | **p = 0.00202**  **q = 0.00303** |
| Salience | 846 | -0.005 ± 0.015 | -0.003 ± 0.018 | 0.009 ± 0.027 | p = 0.206  q = 0.206 | **p = 1.75e-11 q = 5.25e-11** | **p = 1.85e-05**  **q = 2.78e-05** |
| Subcortical | 853 | -0.005 ± 0.015 | -0.002 ± 0.021 | 0.008 ± 0.027 | **p = 0.00687**  **q = 0.0103** | **p = 9.59e-11 q = 2.88e-10** | **p = 0.0164**  **q = 0.0164** |
| Cerebellar | 882 | -0.006 ± 0.02 | 0.001 ± 0.021 | 0.006 ± 0.032 | **p = 0.00384**  **q = 0.00576** | **p = 3.77e-08 q = 1.13e-07** | p = 0.104  q = 0.104 |

**Table S20.** *Low-frequency spectral entropy (lfSE, 0.01-0.045 Hz) value distributions by whole-brain, network and clinical group*. Reported values have had covariates linearly regressed and undergone site harmonization. n=sample size. The medial frontal, default mode, motor, visual association, salience, and subcortical networks were harmonized using the mean only algorithm.

|  |  | Clinical Groups (median +/- std) | | | | Wilcoxon Rank Pairwise p | | |
| --- | --- | --- | --- | --- | --- | --- | --- | --- |
| Scale | n | Controls | Mood | Psychosis | | C vs M | C vs P | M vs P |
| Whole-brain | 823 | -0.005 ± 0.014 | -0.002 ± 0.018 | 0.007 ± 0.023 | p = 0.061  q = 0.061 | | **p = 2.84e-12 q = 8.52e-12** | **p = 0.00107**  **q = 0.0016** |
| Medial Frontal | 846 | -0.006 ± 0.014 | -0.002 ± 0.014 | 0.009 ± 0.027 | **p = 0.00439**  **q = 0.00439** | | **p = 1.12e-13 q = 3.36e-13** | **p = 0.00212**  **q = 0.00318** |
| Frontoparietal | 853 | -0.005 ± 0.013 | -0.001 ± 0.017 | 0.007 ± 0.023 | **p = 0.00096 q = 0.00144** | | **p = 9.96e-11 q = 2.99e-10** | **p = 0.0338**  **q = 0.0338** |
| Default Mode | 882 | -0.005 ± 0.014 | -0.003 ± 0.018 | 0.008 ± 0.029 | **p = 0.00446 q = 0.00668** | | **p = 1.63e-10, q = 4.9e-10** | **p = 0.0126**  **q = 0.0126** |
| Motor | 881 | -0.006 ± 0.013 | -0.003 ± 0.018 | 0.008 ± 0.026 | **p = 0.00158, q = 0.00238** | | **p = 2.36e-11, q = 7.09e-11** | **p = 0.0304**  **q = 0.0304** |
| Visual I | 1082 | -0.002 ± 0.017 | -0.008 ± 0.031 | 0.009 ± 0.026 | p = 0.822  q = 0.822 | | **p = 1.7e-09, q = 5.11e-09** | **p = 2.05e-06**  **q = 3.07e-06** |
| Visual II | 954 | -0.002 ± 0.017 | -0.005 ± 0.025 | 0.007 ± 0.026 | p = 0.75  q = 0.75 | | **p = 1.15e-05, q = 3.44e-05** | **p = 0.000121 q = 0.000181** |
| Visual Association | 906 | -0.005 ± 0.015 | -0.006 ± 0.023 | 0.01 ± 0.028 | p = 0.186  q = 0.186 | | **p = 1.37e-12, q = 4.12e-12** | **p = 4.51e-05**  **q = 6.77e-05** |
| Salience | 846 | -0.005 ± 0.013 | -0.003 ± 0.02 | 0.009 ± 0.027 | **p = 0.000993**  **q = 0.00149** | | **p = 5.21e-09, q = 1.56e-08** | p = 0.0706  q = 0.0706 |
| Subcortical | 853 | -0.005 ± 0.014 | -0.005 ± 0.022 | 0.009 ± 0.026 | p = 0.0528  q = 0.0528 | | **p = 4.53e-11, q = 1.36e-10** | **p = 0.00197**  **q = 0.00296** |
| Cerebellar | 882 | -0.006 ± 0.017 | 0 ± 0.02 | 0.006 ± 0.033 | **p = 0.000995**  **q = 0.00149** | | **p = 1.3e-06, q = 3.91e-06** | p = 0.39  q = 0.39 |

**Table S21.** *Low-frequency spectral entropy (lfSE, 0.035-0.090 Hz) value distributions by whole-brain, network and clinical group*. Reported values have had covariates linearly regressed and undergone site harmonization. n=sample size. The medial frontal, default mode, motor, visual association, salience, and subcortical networks were harmonized using the mean only algorithm.

| Scale | Sample Size | 0.01-0.045 Hz | 0.035-0.090 |
| --- | --- | --- | --- |
| **Whole-brain** | **32** | **0.35 (p = 0.0471*)** | **-0.41 (p = 0.0182*)** |
| **Whole-brain with Available Nodes** | **62** | **0.36 (p = 0.00455*)** | **-0.36 (p = 0.00421*)** |
| Medial Frontal | 48 | 0.11 (p = 0.453*, q = 0.647) | -0.11 (p = 0.467*, q = 0.655) |
| Frontoparietal | 41 | -0.01 (p = 0.934*, q = 0.934) | -0.03 (p = 0.849*, q = 0.849) |
| Default Mode | 58 | 0.17 (p = 0.199*, q = 0.572) | -0.13 (p = 0.317*, q = 0.627) |
| Motor | 35 | -0.25 (p = 0.154*, q = 0.572) | 0.32 (p = 0.0617*, q = 0.335) |
| Visual I | 57 | 0.15 (p = 0.252*, q = 0.572) | -0.22 (p = 0.1*, q = 0.335) |
| Visual II | 56 | 0.05 (p = 0.719*, q = 0.898) | -0.14 (p = 0.304*, q = 0.627) |
| Visual Association | 44 | -0.03 (p = 0.865*, q = 0.934) | -0.09 (p = 0.552*, q = 0.655) |
| Salience | 49 | 0.14 (p = 0.34*, q = 0.572) | -0.08 (p = 0.589*, q = 0.655) |
| Subcortical | 56 | 0.13 (p = 0.343*, q = 0.572) | -0.12 (p = 0.376*, q = 0.627) |
| Cerebellar | 32 | -0.24 (p = 0.192*, q = 0.572) | 0.33 (p = 0.0671*, q = 0.335) |

**Table S22***.* *Frequency-band sensitivity analysis for BDI-II associations with ALFF.* Associations between BDI-II scores and ALFF filtered at distinct low-frequency spectral bands across scales of analysis. The medial frontal, default mode, motor, visual association, salience, and subcortical networks were harmonized using the mean only algorithm.

| **Scale** | **0.01-0.045 Hz** | **0.035-0.090** |
| --- | --- | --- |
| Whole-brain | -0.10 (p=0.1301*) | -0.03 (p=0.6375*) |
| Medial Frontal | -0.12 (p=0.0680*, q=0.2260) | 0.03 (p=0.6210*, q=0.8340) |
| Frontoparietal | -0.11 (p=0.0970*, q=0.2430) | -0.04 (p=0.5230*, q=0.8340) |
| Default Mode | -0.08 (p=0.2280*, q=0.3670) | 0.02 (p= 0.7510*, q=0.8340) |
| Motor | -0.07 (p=0.2930*, q=0.3670) | -0.05 (p=0.4710*, q=0.8340) |
| Visual I | -0.15 (p=0.0210*, q=0.2120) | 0.03 (p=0.6850*, q=0.8340) |
| Visual II | -0.08 (p=0.2340*, q=0.3670) | -0.09 (p=0.1920*, q=0.8340) |
| Visual Association | -0.13 (p=0.0550*, q=0.2260) | -0.05 (p=0.4230*, q=0.8340) |
| Salience | -0.07 (p=0.2900*, q=0.3670) | -0.03 (p=0.6460*, q=0.8340) |
| Subcortical | -0.03 (p=0.6410*, q=0.6410) | -0.06 (p=0.3700*, q=0.8340) |
| Cerebellar | -0.06 (p=0.3970*, q=0.4420) | 0.004 (p=0.9480*, q=0.9480) |

**Table S23***.* *Frequency-band sensitivity analysis for positive PANSS associations with ALFF*. Associations between positive PANSS scores and ALFF filtered at distinct low-frequency spectral bands across scales of analysis. The medial frontal, default mode, motor, visual association, salience, and subcortical networks were harmonized using the mean only algorithm.

| **Scale** | Sample Size | 0.01-0.045 Hz | 0.035-0.090 |
| --- | --- | --- | --- |
| **Whole-brain** | **32** | **-0.42 (p = 0.0172*)** | **-0.51 (p = 0.00271*)** |
| **Whole-brain with Available Nodes** | **62** | **-0.32 (p = 0.0116*)** | **-0.35 (p = 0.00514*)** |
| Medial Frontal | 48 | -0.19 (p = 0.209*, q = 0.396) | -0.21 (p = 0.174*, q = 0.36) |
| Frontoparietal | 41 | 0.02 (p = 0.912*, q = 0.912) | -0.08 (p = 0.626*, q = 0.626) |
| Default Mode | 58 | -0.2 (p = 0.144*, q = 0.396) | -0.23 (p = 0.0865*, q = 0.308) |
| Motor | 35 | 0.39 (p = 0.0245*, q = 0.245) | 0.18 (p = 0.322*, q = 0.41) |
| Visual I | 57 | -0.13 (p = 0.351*, q = 0.396) | -0.18 (p = 0.18*, q = 0.36) |
| Visual II | 56 | -0.15 (p = 0.291*, q = 0.396) | -0.26 (p = 0.0653*, q = 0.308) |
| Visual Association | 44 | -0.17 (p = 0.291*, q = 0.396) | -0.15 (p = 0.328*, q = 0.41) |
| Salience | 49 | -0.19 (p = 0.207*, q = 0.396) | -0.17 (p = 0.258*, q = 0.41) |
| Subcortical | 56 | -0.14 (p = 0.31*, q = 0.396) | -0.23 (p = 0.0924*, q = 0.308) |
| Cerebellar | 32 | 0.17 (p = 0.357*, q = 0.396) | 0.11 (p = 0.578*, q = 0.626) |

**Table S24***. Frequency-band sensitivity analysis for BDI-II associations with lfSE.* Associations between BDI-II scores and lfSE filtered at distinct low-frequency spectral bands across scales of analysis. Bolded table entries represent significant comparisons. The medial frontal, default mode, motor, visual association, salience, and subcortical networks were harmonized using the mean only algorithm.

| **Scale** | **0.01-0.045 Hz** | **0.035-0.090 Hz** |
| --- | --- | --- |
| **Whole-brain** | **-0.14 (p=0.03321*)** | **-0.15 (p=0.02674*)** |
| Medial Frontal | -0.10 (p=0.1130*, q=0.1250) | -0.13 (p=0.0570*, q=0.0820) |
| Frontoparietal | -0.15 (p= 0.0250*, q=0.0640) | -0.14 (p=0.0310*, q=0.0820) |
| Default Mode | -0.12 (p=0.0700*, q=0.0890) | -0.16 (p=0.0150*, q=0.0750) |
| Motor | -0.16 (p=0.0150*, q=0.0640) | -0.13 (p=0.0530*, q=0.0820) |
| Visual I | -0.09 (p=0.1810*, q=0.1810) | -0.13 (p=0.0510*, q=0.0820) |
| Visual II | -0.15 (p=0.0230*, q=0.0640) | -0.11 (p=0.1010*, q=0.1010) |
| Visual Association | -0.12 (p=0.0640*, q=0.0890) | -0.13 (p=0.0470*, q=0.0820) |
| Salience | -0.12 (p= 0.0710*, q=0.0890) | -0.12 (p=0.0700*, q=0.0880) |
| Subcortical | -0.12 (p= 0.0650*, q=0.0890) | -0.11 (p=0.0840*, q= 0.0930) |
| **Cerebellar** | **-0.20 (p=0.0020*, q=0.020)** | **-0.22 (p=0.0010*, q= 0.0060)** |

**Table S25***.* *Frequency-band sensitivity analysis for positive PANSS associations with lfSE*. Associations between positive PANSS scores and lfSE filtered at distinct low-frequency spectral bands across scales of analysis. Bolded table entries represent significant comparisons. The medial frontal, default mode, motor, visual association, salience, and subcortical networks were harmonized using the mean only algorithm.

| **Scale** | n | Clinical Groups (median +/- std) | | | Wilcoxon Rank Pairwise p | | |
| --- | --- | --- | --- | --- | --- | --- | --- |
|  |  | C | M | P | C vs M | C vs P | M vs P |
| Whole-brain | 554 (C=232 M=105  P=217) | 0.009 ± 0.012 | -0.016 ± 0.025 | -0.002 ± 0.027 | **p = 8.87e-19**  **q = 2.66e-18** | **p = 1.72e-11 q = 2.58e-11** | **p = 0.000547 q = 0.000547** |
| Medial Frontal | 554  (C=232 M=105  P=217) | 0.009 ± 0.015 | -0.017 ± 0.026 | -0.001 ± 0.028 | **p = 3.84e-19**  **q = 1.15e-18** | **p = 8.48e-09 q = 1.27e-08** | **p = 0.000108 q = 0.000108** |
| Frontoparietal | 554  (C=232 M=105  P=217) | 0.009 ± 0.014 | -0.017 ± 0.026 | -0.002 ± 0.029 | **p = 7.27e-17**  **q = 2.18e-16** | **p = 4.95e-08 q = 7.43e-08** | **p = 0.000214 q = 0.000214** |
| Default Mode | 554  (C=232 M=105  P=217) | 0.008 ± 0.015 | -0.014 ± 0.028 | -0.002 ± 0.031 | **p = 5.57e-12**  **q = 1.67e-11** | **p = 1.63e-07 q = 2.45e-07** | **p = 0.00577**  **q = 0.00577** |
| Motor | 555  (C=232 M=106  P=217) | 0.009 ± 0.015 | -0.016 ± 0.025 | -0.001 ± 0.028 | **p = 3.79e-17 q = 1.14e-16** | **p = 7.74e-09 q = 1.16e-08** | **p = 0.000474 q = 0.000474** |
| Visual I | 558  (C=232 M=109,  P=217) | 0.009 ± 0.017 | -0.017 ± 0.028 | -0.001 ± 0.029 | **p = 2.08e-15 q = 6.25e-15** | **p = 9.7e-07**  **q = 1.45e-06** | **p = 7.91e-05**  **q = 7.91e-05** |
| Visual II | 556  (C=232 M=107  P=217) | 0.009 ± 0.019 | -0.017 ± 0.028 | -0.001 ± 0.03 | **p = 5.86e-16 q = 1.76e-15** | **p = 1e-05**  **q = 1.5e-05** | **p = 3.27e-05**  **q = 3.27e-05** |
| Visual Association | 555  (C=232 M=106  P=217) | 0.009 ± 0.016 | -0.017 ± 0.028 | -0.001 ± 0.028 | **p = 2.27e-16 q = 6.81e-16** | **p = 6.25e-08 q = 9.37e-08** | **p = 8.26e-05**  **q = 8.26e-05** |
| Salience | 555  (C=232 M=106  P=217) | 0.009 ± 0.014 | -0.016 ± 0.025 | -0.002 ± 0.028 | **p = 3.67e-17 q = 1.1e-16** | **p = 1.77e-09 q = 2.65e-09** | **p = 0.000389**  **q = 0.000389** |
| Subcortical | 554  (C=232 M=105  P=217) | 0.009 ± 0.013 | -0.015 ± 0.025 | -0.002 ± 0.027 | **p = 6.9e-16, q = 2.07e-15** | **p = 1e-10**  **q = 1.5e-10** | **p = 0.0017**  **q = 0.0017** |
| Cerebellar | 554  (C=232 M=105  P=217) | 0.008 ± 0.014 | -0.012 ± 0.027 | -0.004 ± 0.04 | **p = 3.7e-10, q = 1.11e-09** | **p = 5.42e-05 q = 8.13e-05** | **p = 0.0169**  **q = 0.0169** |

**Table S26.** *Effect of Medication Exposure on lfSE Group Differences.* A subsample of the patient group had known exposure to a psychiatric medication and a sensitivity analysis was performed on this subsample. Significant group differences for case-case and case-control conditions were identified. Motor and visual I networks were analyzed using the mean only harmonization algorithm (see Supplementary methods).

**Supplementary References**

1.	COBRE Phase 3 | The Mind Research Network (MRN). Accessed February 14, 2025. https://www.mrn.org/common/cobre-phase-3

2.	Lewandowski KE, Bouix S, Ongur D, Shenton ME. Neuroprogression across the Early Course of Psychosis. *J Psychiatr Brain Sci*. 2020;5:e200002. doi:10.20900/jpbs.20200002

3.	American Psychiatric Association. *Diagnostic and Statistical Manual of Mental Disorders*. Fifth Edition. American Psychiatric Association; 2013. Accessed March 26, 2025. https://psychiatryonline.org/doi/book/10.1176/appi.books.9780890425596

4.	Tanaka SC, Yamashita A, Yahata N, et al. A multi-site, multi-disorder resting-state magnetic resonance image database. *Sci Data*. 2021;8(1):227. doi:10.1038/s41597-021-01004-8

5.	Poldrack RA, Congdon E, Triplett W, et al. A phenome-wide examination of neural and cognitive function. *Sci Data*. 2016;3(1):160110. doi:10.1038/sdata.2016.110
